# ClinAgent: A Five-Layer Architecture for Autonomous Clinical Trial Statistical Programming

**DOI:** 10.64898/2026.01.09.26343542

**Authors:** Jaime Yan

## Abstract

Clinical trial statistical programming requires 12-24 FTE-months for a typical Phase 3 study, producing 100-500 tables, listings, and figures (TLFs) across 8-15 ADaM domains. Modern AI coding agents (Augment Code, Claude Code, Cline, Cursor) demonstrate remarkable reasoning capabilities but lack the domain-specific tools needed for clinical programming: they cannot read SAS datasets, parse ADaM specifications, analyze regulatory-relevant log issues, or generate CDISC-compliant code without extensive guidance.

We present **ClinAgent**, a **skill and tool layer** that augments any AI coding agent with clinical programming capabilities. Unlike approaches that build custom LLM agents, ClinAgent provides domain expertise as MCP (Model Context Protocol) tools that any compatible agent can invoke. Our “Thin MCP, Thick Skills” design separates minimal data access operations from rich domain logic: Skills package embedded prompts, rule engines, and decision trees encoding expert knowledge; MCP tools provide stateless I/O for SAS datasets, Excel specifications, and log files. Users retain their preferred AI agent while gaining clinical programming tools.

We validate ClinAgent’s nine skills (SK-001 Study Setup through SK-009 eSub Packaging) using artifacts from a production Phase 2 cardiovascular study: specifications, SAS programs, and synthetic datasets matching the study structure (11 ADaM domains, 93,239 synthetic observations). All skills pass functional validation. Deterministic components achieve high accuracy: log analysis identifies 1 error and 7 warnings across 10 logs with 100% precision; data validation matches all 56 ADSL variables. Prompt-based specification generation achieves 72.1% derivation accuracy overall, with simple domains exceeding 96%. Our contributions include: (1) an agent-augmentation architecture enabling any AI coding agent to perform clinical programming; (2) nine production-ready skills with embedded prompts and deterministic rule engines; (3) MCP tool implementations for clinical data formats; (4) a validation methodology distinguishing tool correctness from LLM performance; and (5) quantitative results from production study evaluation.

**Availability:** Architecture specifications at https://github.com/yanmingyu92/ClinAgent.

## 1 Introduction

### 1.1 The Clinical Programming Bottleneck

The pharmaceutical industry invests over $2.6 billion and 10-15 years to bring a single drug to market [1]. Within this process, clinical trial statistical programming—the conversion of raw clinical data into analysis datasets, tables, listings, and figures (TLFs)—represents a critical path activity that directly impacts regulatory submission timelines. A typical Phase 3 clinical trial requires:

- 8-15 Analysis Data Model (ADaM) domains (ADSL, ADAE, ADLB, etc.)
- 100-500 TLFs for regulatory submission
- 12-24 FTE-months of programming effort [2]
- 30-50% quality control overhead

Despite significant advances in clinical data standards (CDISC) and statistical computing, the programming workflow remains largely manual, dependent on experienced SAS programmers who are increasingly scarce and expensive. Industry surveys report 15-25% rework rates due to specification changes and quality issues [2].

### 1.2 The Rise of AI Coding Agents

A new generation of AI coding agents—including Augment Code, Claude Code (Anthropic), Cline, Cursor, and GitHub Copilot—has emerged that goes beyond simple code completion. These agents can:

- Understand natural language task descriptions
- Read and write files autonomously
- Execute code and interpret results
- Iterate based on errors and feedback

However, these general-purpose agents lack the **domain-specific tools** needed for clinical programming. They cannot read SAS datasets, parse ADaM specifications, analyze SAS logs for regulatory-relevant issues, or generate CDISC-compliant code without extensive manual guidance.

### 1.3 Research Gap: The Missing Tool Layer

The gap is not in AI reasoning capability—modern agents are remarkably capable. The gap is in **domain-specific tooling**:

- **No clinical data tools**: Agents cannot natively read SAS7BDAT files, Excel specifications, or RTF outputs
- **No domain knowledge packaging**: CDISC standards, ADaM derivation patterns, and QC rules are not accessible to agents
- **No workflow orchestration**: The end-to-end pipeline (spec → code → execute → validate → package) requires coordinated tool sequences
- **No compliance infrastructure**: GxP audit trails and data masking are not built into general-purpose agents

### 1.4 Our Contribution: A Skill and Tool Layer for Clinical Programming

We present **ClinAgent**, a **skill and tool layer** designed to augment any AI coding agent with clinical programming capabilities. Crucially, ClinAgent is *not* an LLM or agent itself—it is a collection of:

1. **Skills**: Self-contained packages combining domain knowledge (CDISC standards, SAS patterns), embedded prompts (guiding agent reasoning), rule engines (deterministic validation logic), and tool bindings.
2. **MCP Tools**: Model Context Protocol [3] servers providing standardized data access (read SAS datasets, parse Excel specs, analyze logs) that any MCP-compatible agent can invoke.
3. **Compliance Infrastructure**: Audit logging, data masking, and access control ensuring GxP-ready operation.

This design embodies the principle of **“Thin MCP, Thick Skills”** : data access tools (MCP) are minimal and stateless, while domain logic is packaged into rich, testable skill definitions.

### 1.5 Key Insight: Prompts as Packaged Expertise

A central innovation is treating **prompts as first-class artifacts** within skills. Rather than requiring users to craft prompts for each task, ClinAgent skills contain:

- **Task-specific prompts**: Pre-engineered instructions for ADaM coding, log review, spec interpretation
- **Few-shot examples**: Curated input-output pairs demonstrating correct behavior
- **Constraint specifications**: Rules the agent must follow (CDISC compliance, naming conventions)
- **Decision trees**: Navigation logic for complex multi-step reasoning

When a user asks their agent (Augment, Claude Code, etc.) to “review the ADSL log for errors,” the agent invokes ClinAgent’s SK-005 skill, which provides both the analytical prompt *and* the MCP tools to read the log file. The agent handles reasoning; ClinAgent provides domain expertise.

### 1.6 Contributions

1. **Agent-Augmentation Architecture**: A five-layer design where ClinAgent provides Skills and Tools while users retain their choice of AI agent, enabling domain capability without agent lock-in.
2. **Nine Production-Ready Skills**: SK-001 (Study Setup) through SK-009 (eSub Packaging) covering the complete clinical programming workflow, each with embedded prompts, rules, and tool bindings.
3. **MCP Tool Suite**: Six Model Context Protocol servers for clinical data operations (SAS datasets, Excel specs, logs, RTF comparison, folder management).
4. **Validation Framework**: Methodology for evaluating skill correctness independent of agent choice, with results using production study artifacts (specifications, programs, and synthetic data across 11 ADaM domains).
5. **Open Design**: Architecture specifications enabling community extension and adoption across organizations.

### 1.7 Paper Organization

Section 2 reviews clinical programming workflows and the Model Context Protocol. Section 3 presents our agent-augmentation architecture. Section 4 describes the skill library. Section 5 defines our skill validation methodology. Section 6 reports results from production study evaluation. Section 7 discusses implications, and Section 8 concludes.

## 2 Background and Related Work

### 2.1 Clinical Trial Statistical Programming

Clinical trial statistical programming transforms raw clinical data into regulatory-compliant analysis datasets and outputs. The workflow follows CDISC (Clinical Data Interchange Standards Consortium) standards:

**Study Data Tabulation Model (SDTM)**: Standardized format for raw clinical data organized by domains (Demographics, Adverse Events, Laboratory, etc.).

**Analysis Data Model (ADaM)**: Derived analysis-ready datasets built from SDTM, following implementation guides for subject-level (ADSL), Basic Data Structure (BDS), and occurrence (OCCDS) datasets.

**Tables, Listings, and Figures (TLFs)**: Statistical outputs generated from ADaM datasets for clinical study reports and regulatory submissions.

The typical workflow proceeds as:

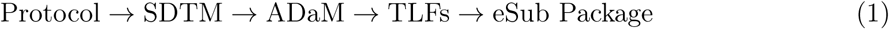

Each stage requires specification development, code writing, execution, quality control (QC), and validation—all subject to Good Clinical Practice (GxP) requirements.

### 2.2 LLM Agent Architectures

Recent advances in LLM-based agents have produced several architectural patterns:

**ReAct (Reasoning + Acting)** [4]: Interleaves reasoning traces with tool use, enabling multi-step problem solving.

**Tool-Augmented LLMs**: Systems like Toolformer [5] and GPT-4 function calling enable LLMs to invoke external tools.

**Multi-Agent Systems**: Frameworks like AutoGen [6] and CrewAI orchestrate multiple specialized agents for complex tasks.

**Model Context Protocol (MCP)** [3]: Open protocol standardizing how LLMs connect to data sources and tools through JSON-RPC interfaces.

These architectures share common components:

- **Orchestration Layer**: Task planning and coordination
- **Tool Layer**: External capability access
- **Memory Layer**: Context persistence
- **Execution Layer**: Action implementation

### 2.3 AI in Clinical Research

AI applications in clinical research have primarily focused on:

**Clinical Trial Design**: ML-based patient selection, site identification, and protocol optimization [7].

**Data Management**: Automated data cleaning, anomaly detection, and query generation.

**Medical Coding**: NLP for adverse event coding (MedDRA) and medication coding (WHO Drug).

**Document Generation**: LLM-assisted clinical study report writing.

### 2.4 Recent LLM Applications in Clinical Trials

Recent work has begun applying LLMs to specific clinical trial tasks:

**Trial Matching and Eligibility Screening**: Beattie et al. [13] demonstrated LLM-based clinical trial matching achieving 98.6% accuracy on eligibility determination and 38.1% triage success rate. These systems focus on patient-trial matching rather than statistical programming.

**CDISC-Compliant Specification Tools**: Chen et al. [14] developed PmWebSpec, a web application for creating CDISC-compliant pharmacometric analysis dataset specifications. While semi-automated, it requires manual data entry and does not integrate with AI agents.

**Cross-Study Analysis Packages**: Snyder et al. [15] presented sendigR, an R package for SEND (Standard for Exchange of Nonclinical Data) cross-study analysis. This addresses nonclinical data only and is rule-based rather than AI-driven.

**Multi-Agent Knowledge Engines**: Vieira-Vieira et al. [16] developed PRINCE, a multi-agent knowledge engine for preclinical drug development. While demonstrating multi-agent orchestration, it focuses on knowledge retrieval rather than code generation.

**Regulatory Report Generation**: Emerging work on RAG (Retrieval-Augmented Generation) systems for clinical study report drafting reports 75% time reduction in document preparation [17]. However, these systems generate narrative text, not executable code.

**CDISC Analysis Results Standards**: The CDISC Analysis Results Standard (ARS) [18] provides a metadata-driven approach to TLF specification but requires implementation tooling—a gap ClinAgent addresses.

### 2.5 Gap in Statistical Programming Automation

Despite these advances, *statistical programming automation* remains underexplored. Ravula [19] notes that while R, Python, and AI integration show promise, validated end-to-end pipelines for submission-grade outputs do not exist. Existing partial solutions include:

- Rule-based macro libraries (company-specific, not AI-driven)
- Template-based code generation (limited flexibility)
- Copilot-style code completion (no workflow awareness)
- Web-based specification tools (manual, not agent-integrated)

**Key Gap**: No existing system provides (1) AI agent integration via standard protocols, (2) end-to-end workflow from specification to eSub, and (3) validated tools for submission-grade outputs. ClinAgent addresses this gap.

### 2.6 Gap Analysis

Table 1 compares existing approaches with our framework:

**Table 1:**
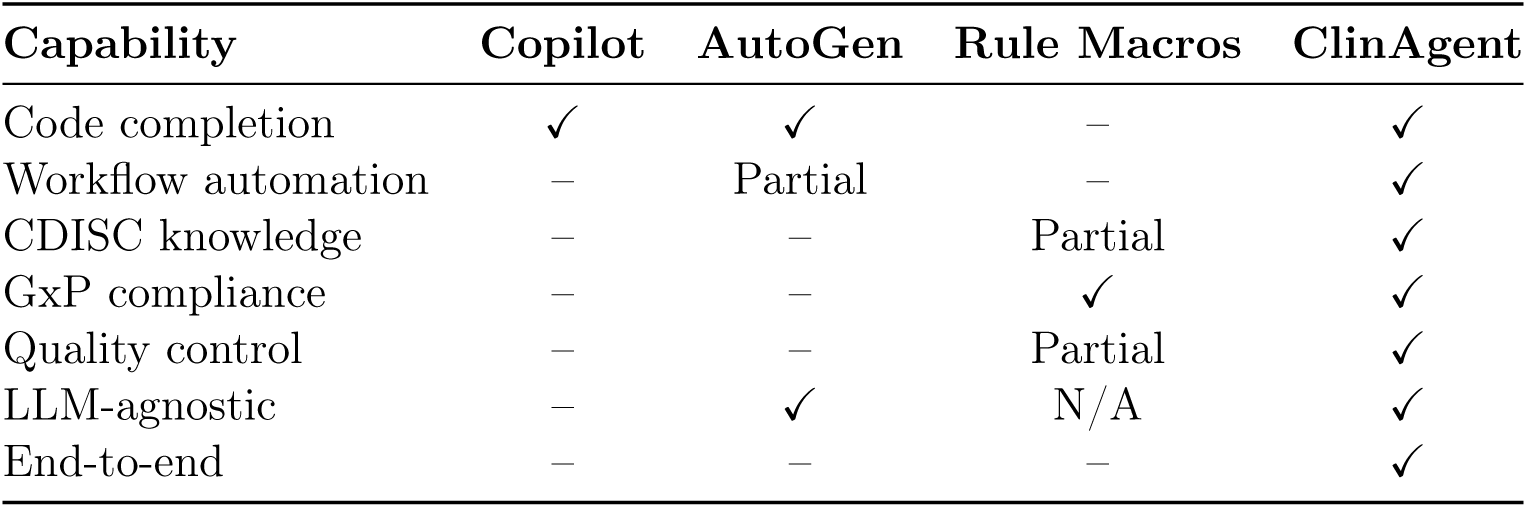
Comparison of AI approaches for clinical programming.

Our framework addresses the unique intersection of:

1. LLM agent capabilities (reasoning, planning, tool use)
2. Domain-specific knowledge (CDISC, ADaM, macros)
3. Regulatory requirements (audit trails, validation)
4. Production engineering (reliability, determinism)

## 3 Architecture Design

### 3.1 Core Concept: Agent Augmentation, Not Agent Replacement

ClinAgent is **not** an AI agent or LLM system. It is a **tool and skill layer** that augments existing AI coding agents with clinical programming capabilities. This distinction is fundamental:

**Table 2:** ClinAgent vs. Traditional Agent Frameworks.

| Aspect | Traditional (AutoGPT, LangChain) | ClinAgent |
| --- | --- | --- |
| LLM integration | Built-in, specific model | None—uses user’s agent |
| Agent logic | Framework provides | User’s agent provides |
| Domain tools | Generic | Clinical-specific |
| Prompt engineering | User responsibility | Packaged in skills |
| Portability | Tied to framework | Works with any MCP agent |

### 3.2 Capability Gap: What Generic Agents Cannot Do

Before detailing our design, we make explicit the capability gap that motivates ClinAgent. Table 3 compares what clinical programmers can accomplish with generic AI agents versus agents augmented with ClinAgent.

**Table 3:** Capability Comparison: Generic AI Agents vs. ClinAgent-Augmented Agents.

| Clinical Programming Task | *Copilot* | *Claude* | *Cursor* | *Augment* | *+ClinAgent* |
| --- | --- | --- | --- | --- | --- |
| Read SAS7BDAT datasets natively | ✗ | ✗ | ✗ | ✗ | ✓ |
| Parse ADaM specification Excel | ✗ | ✗ | ✗ | ✗ | ✓ |
| Classify SAS log issues (ERROR/WARN) | △ | △ | △ | △ | ✓ |
| Generate CDISC-compliant code | △ | △ | △ | △ | ✓ |
| Validate ADaM datasets against spec | ✗ | ✗ | ✗ | ✗ | ✓ |
| Compare RTF outputs programmatically | ✗ | ✗ | ✗ | ✗ | ✓ |
| Apply company-specific macro catalog | ✗ | ✗ | ✗ | ✗ | ✓ |
| Create standardized eSub packages | ✗ | ✗ | ✗ | ✗ | ✓ |
| GxP-compliant audit logging | ✗ | ✗ | ✗ | ✗ | ✓ |
| PHI/PII data masking in context | ✗ | ✗ | ✗ | ✗ | ✓ |

✓ = native capability, △ = possible with extensive prompting, ✗ = not possible

Generic agents have powerful reasoning capabilities but lack **domain-specific tools**. They cannot read proprietary file formats (SAS7BDAT), access structured specifications (ADaM Excel templates), or apply organization-specific knowledge (macro catalogs). ClinAgent bridges this gap by providing the missing tool layer.

### 3.3 Design Principles

**Principle 1: Bring Your Own Agent.** Users choose their preferred AI coding agent (Augment Code, Claude Code, Cline, Cursor, etc.). ClinAgent provides the domain-specific tools these agents need but lack.

**Principle 2: Thin MCP, Thick Skills.** The Model Context Protocol (MCP) layer provides minimal, stateless data operations (read file, write file). All domain logic—CDISC rules, derivation patterns, QC logic—resides in Skills. This enables:

- Independent testing of domain logic
- Easy updates to rules without changing data layer
- Reusable tools across different skills

**Principle 3: Prompts as Packaged Expertise.** Rather than expecting users to prompt-engineer clinical tasks, skills contain pre-built prompts, examples, and constraints. The agent receives domain guidance automatically.

**Principle 4: Deterministic Where It Matters.** Agent reasoning is stochastic; that’s acceptable for creative tasks. But error classification, validation rules, and compliance checks must be deterministic. Skills encode these as rule engines, not LLM calls.

### 3.4 Design Rationale: Why These Architectural Choices?

#### 3.4.1 Why Agent-Augmentation Instead of a Custom Agent?

We could have built a complete clinical programming agent from scratch. We deliberately chose not to, for three reasons:

1. **Rapid Agent Evolution**: General-purpose AI agents (Augment, Claude Code, Cursor) improve monthly. Building our own agent would require us to continuously match these improvements in reasoning, context handling, and user experience—a resource-intensive treadmill.
2. **Enterprise Integration**: Organizations already have preferred development tools and AI assistants. ClinAgent integrates into existing workflows rather than demanding tool replacement, dramatically lowering adoption barriers.
3. **Separation of Concerns**: Agent reasoning and domain tooling are orthogonal concerns. By separating them, we can focus exclusively on clinical programming tools while benefiting from advances in agent capabilities.

The result: when Anthropic improves Claude’s reasoning or Augment adds new IDE features, ClinAgent users benefit automatically—without any framework updates.

#### 3.4.2 Why “Thin MCP, Thick Skills”?

The MCP layer could contain domain logic (e.g., the SAS log parser could classify errors). We deliberately keep MCP “thin” for three reasons:

1. **Testability**: Domain rules embedded in MCP tools require integration testing with actual files. Rules in Skills can be unit-tested with JSON fixtures—faster, more comprehensive, easier to maintain.
2. **Evolvability**: CDISC standards change, company conventions differ, new error patterns emerge. Updating JSON rule files is trivial; updating compiled MCP servers requires deployment cycles.
3. **Transparency**: When rules live in JSON (Skills), domain experts can review and modify them without programming knowledge. When rules are compiled into code (MCP), they become black boxes.

Concrete example: When a new SAS warning pattern appears, we add one line to warning_patterns.json. No code changes, no redeployment, no testing of data access logic.

### 3.5 Five-Layer Architecture

Figure 1 shows how ClinAgent layers interact with external agents.

**Figure 1:**
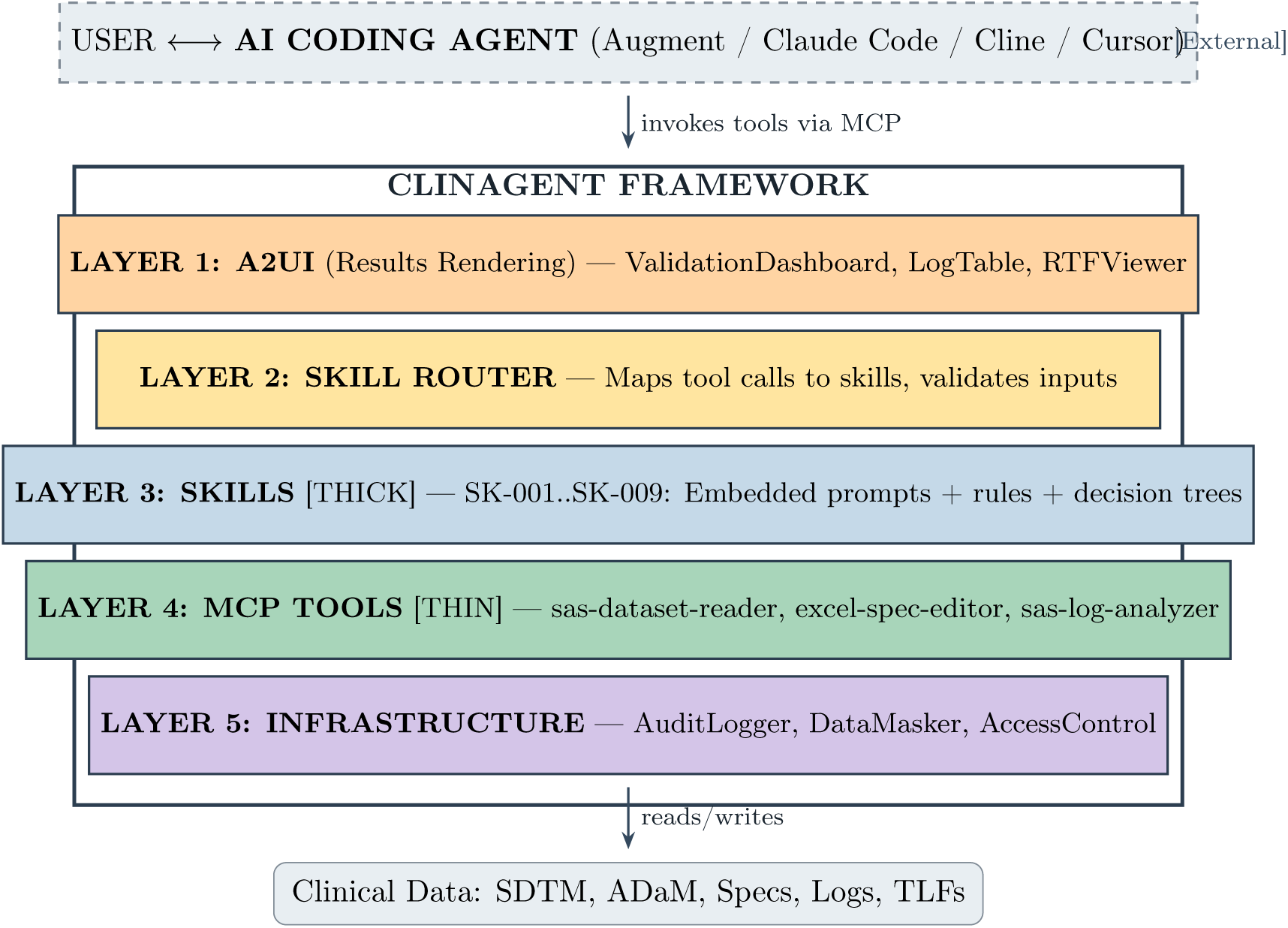
ClinAgent Architecture. The framework provides Skills and MCP Tools that any AI coding agent can invoke. The agent handles reasoning; ClinAgent provides domain expertise and data access.

### 3.6 Layer Details

#### 3.6.1 External: AI Coding Agent (User’s Choice)

The agent is **not part of ClinAgent**. Users work with their preferred agent:

- **Augment Code**: Enterprise-focused, codebase-aware
- **Claude Code**: Anthropic’s coding agent with computer use
- **Cline**: Open-source autonomous coding agent
- **Cursor**: AI-first code editor

These agents connect to ClinAgent via MCP, receiving tools they can invoke and skill-provided prompts that guide their reasoning.

#### 3.6.2 Layer 3: Skills (The “Thick” Layer)

Skills are the core contribution. Each skill is a self-contained package:

**Listing 1:**
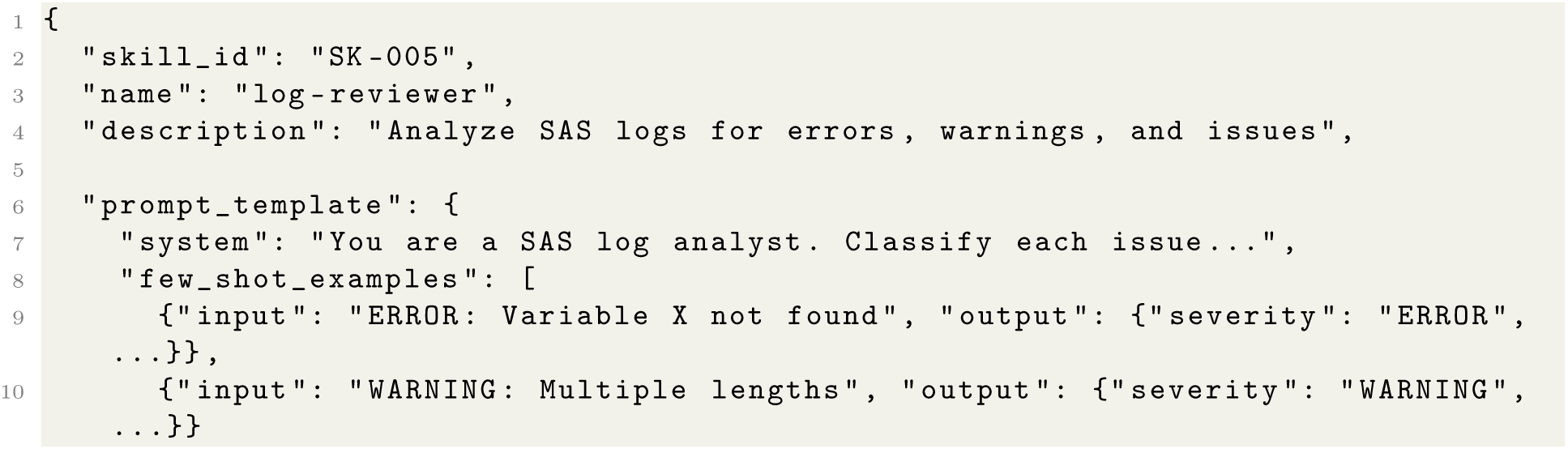
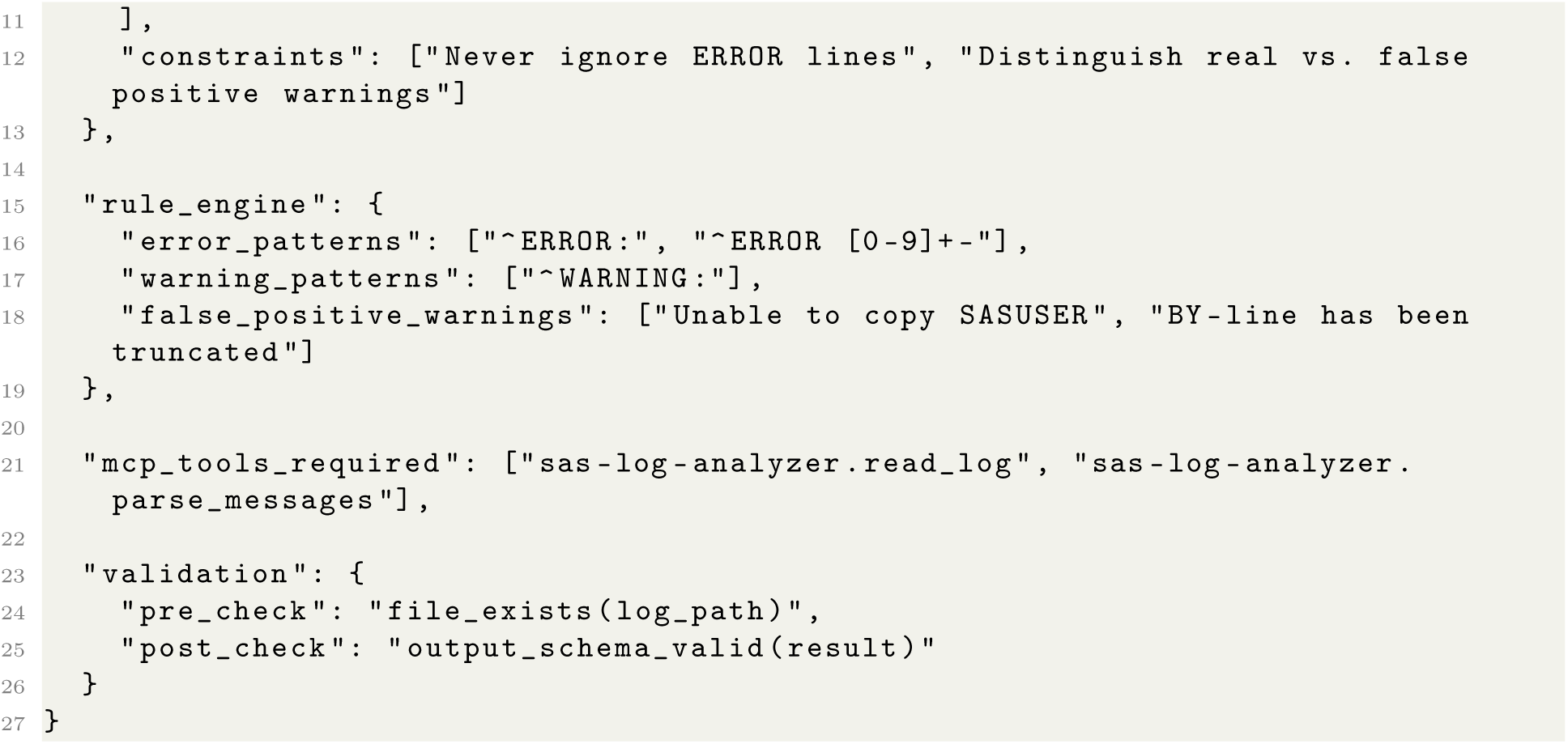
Skill Structure (SK-005 Log Reviewer)

When an agent invokes SK-005, it receives:

1. The prompt_template to guide its reasoning
2. Access to mcp_tools_required for data operations
3. The rule_engine for deterministic classification

The agent reasons about the log content; the skill ensures domain-correct behavior.

#### 3.6.3 Layer 4: MCP Tools (The “Thin” Layer)

MCP servers provide stateless data operations:

**Listing 2:**
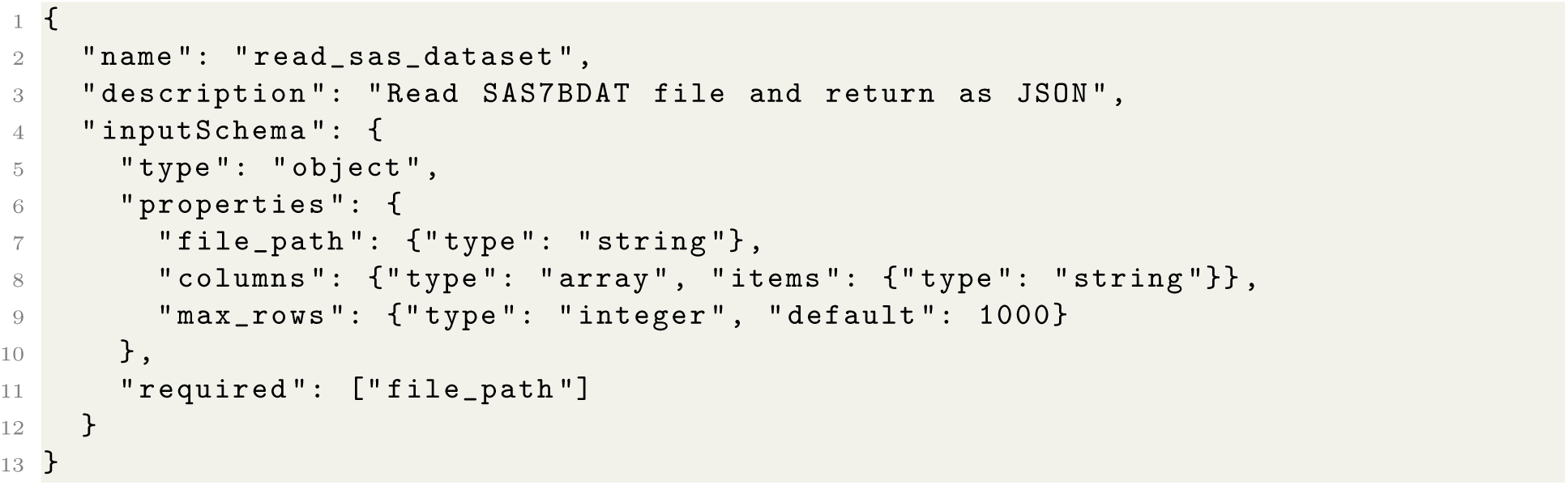
MCP Tool Definition

MCP tools contain **no domain logic**—they don’t know what ADSL means or which variables matter. That knowledge lives in Skills.

#### 3.6.4 Layer 5: Infrastructure (Compliance)

Ensures regulatory readiness:

- **AuditLogger**: Records every tool invocation with timestamps, inputs, outputs
- **DataMasker**: Removes PHI/PII before data reaches agent context
- **AccessControl**: Enforces role-based permissions on sensitive operations
- **ContextMinimizer**: Sends only necessary data to agent, reducing exposure

### 3.7 Concrete Workflow Walkthrough

To make the architecture concrete, we trace a complete interaction from user request to final output.

#### 3.7.1 Scenario: Log Review Request

Consider a user asking their agent: *“Review the ADSL log and summarize any issues.”*

**Figure 2:**
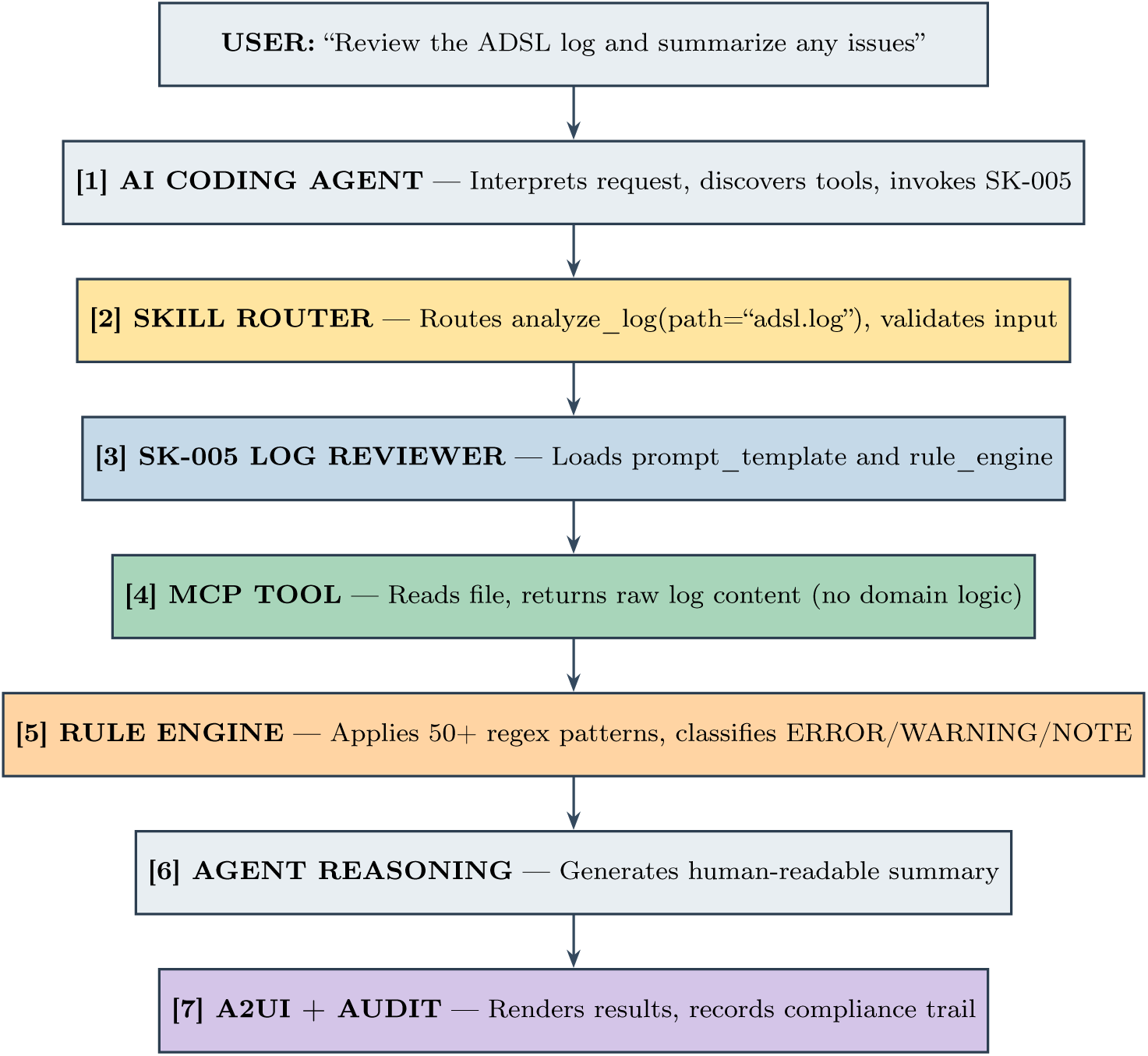
Complete Workflow Trace: Log Review Request. The flow shows how a user request passes through the ClinAgent framework layers.

#### 3.7.2 What Each Component Contributes

**Table 4:** Responsibility Distribution in Log Review Workflow.

| Component | Type | Responsibility |
| --- | --- | --- |
| AI Agent | External | Interpret request, reason, generate summary |
| Skill Router | ClinAgent | Validate input, route to correct skill |
| SK-005 Skill | ClinAgent | Provide domain prompts and rules |
| MCP Tool | ClinAgent | Read file (pure I/O) |
| Rule Engine | ClinAgent | Classify issues deterministically |
| A2UI | ClinAgent | Format output for human review |
| Audit Logger | ClinAgent | Maintain compliance trail |

#### 3.7.3 Concrete Data Flow Example

**Input (Log File Content):**

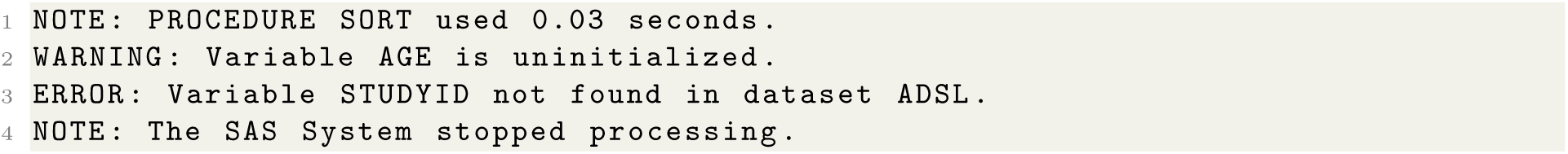

**Rule Engine Output (Deterministic):**

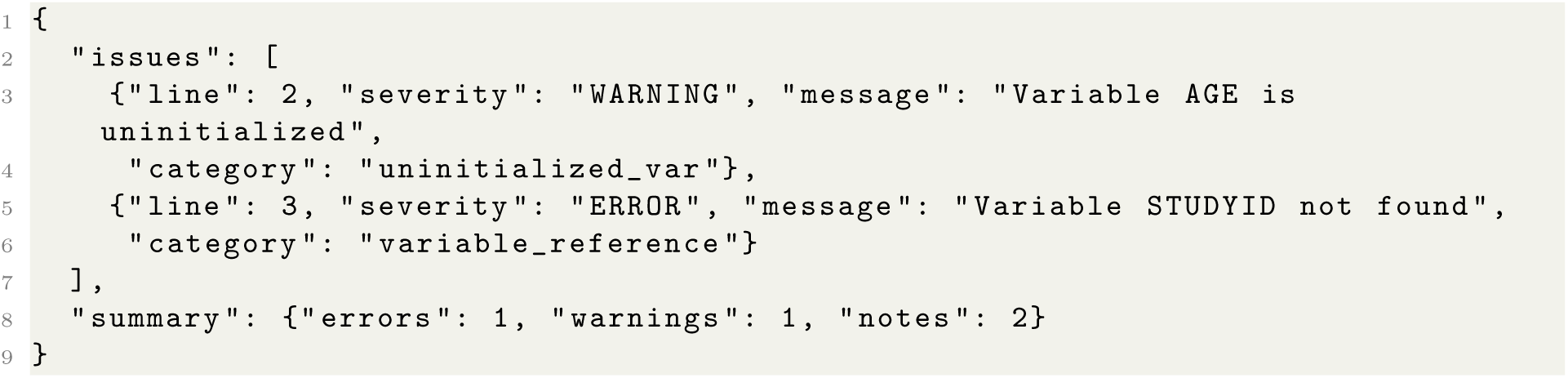

**Agent-Generated Summary (Stochastic):**

*“The ADSL log contains 1 critical error and 1 warning. The ERROR on line 3 indicates STUDYID is missing from the input dataset—check that SDTM DM is being read correctly. The WARNING on line 2 suggests AGE may need initialization before the retain statement.”*

This separation—deterministic classification, stochastic interpretation—ensures reliable issue detection while enabling helpful human-readable explanations.

## 4 Skill Library

### 4.1 Skill Catalog Overview

ClinAgent implements nine production-ready skills covering the complete clinical programming workflow. Table 5 summarizes the skill catalog.

**Table 5:** ClinAgent Skill Catalog.

| ID | Name | Purpose | MCP Tools |
| --- | --- | --- | --- |
| SK-001 | StudySetup | Create folder structure | filesystem |
| SK-002 | SpecEditor | Edit ADaM specifications | excel-spec-editor |
| SK-003 | ADaMCoder | Generate ADaM programs | excel-spec, filesystem |
| SK-004 | SASRunner | Execute SAS programs | process, filesystem |
| SK-005 | LogReviewer | Analyze SAS logs | sas-log-analyzer |
| SK-006 | DataValidator | Validate datasets vs spec | sas-dataset-reader |
| SK-007 | TLFCoder | Generate TLF programs | macro-catalog |
| SK-008 | RTFQC | Compare RTF outputs | rtf-comparator |
| SK-009 | eSubPackager | Create eSub package | filesystem |

### 4.2 Skill Architecture

Each skill follows a standardized structure (Figure 3):

**Figure 3:**
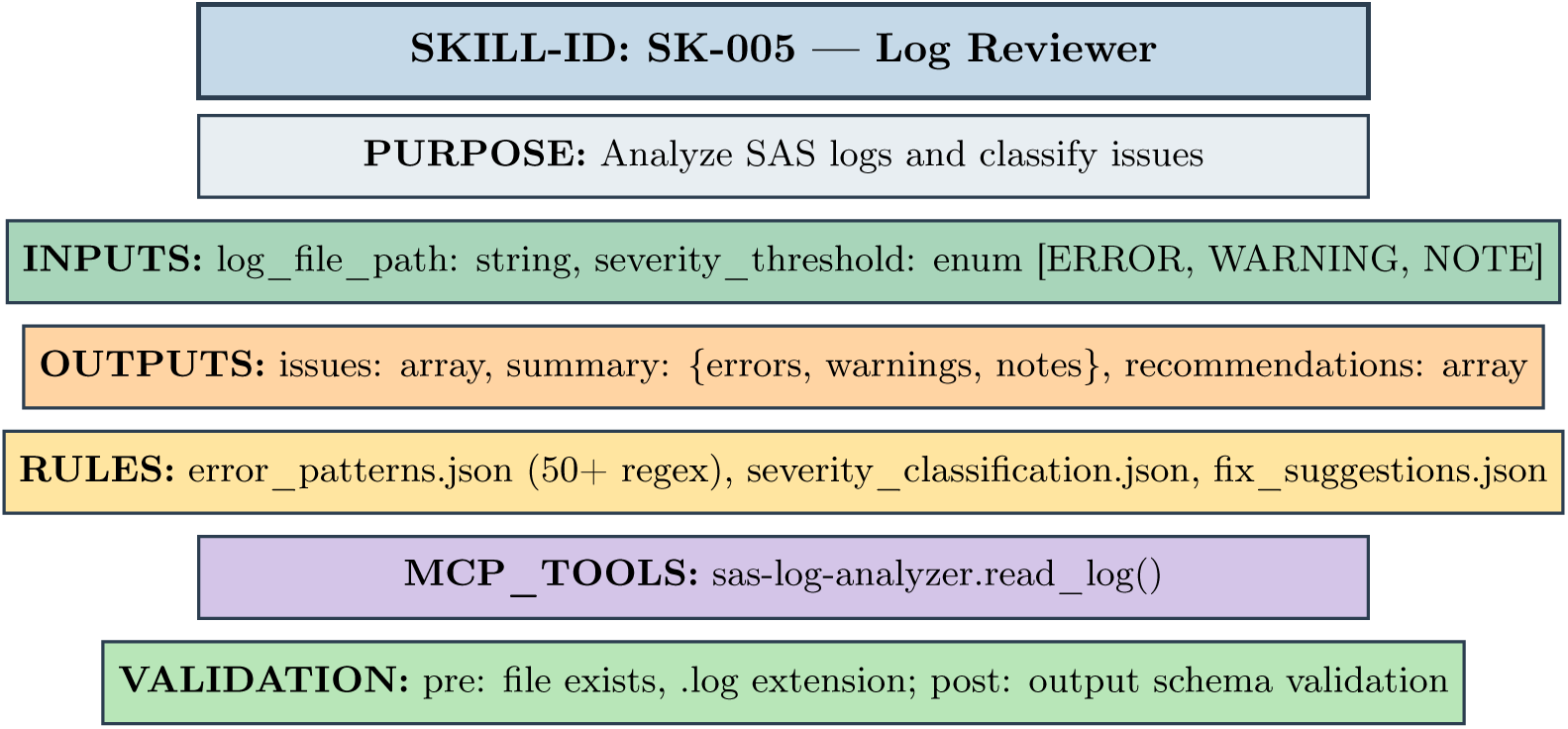
Skill Definition Structure (SK-005 Example). Each skill contains inputs, outputs, rule engines, MCP tool bindings, and validation criteria.

### 4.3 Rule Engines

Skills encode domain expertise in JSON rule engines. Example from SK-005 (Log Reviewer):

**Listing 3:**
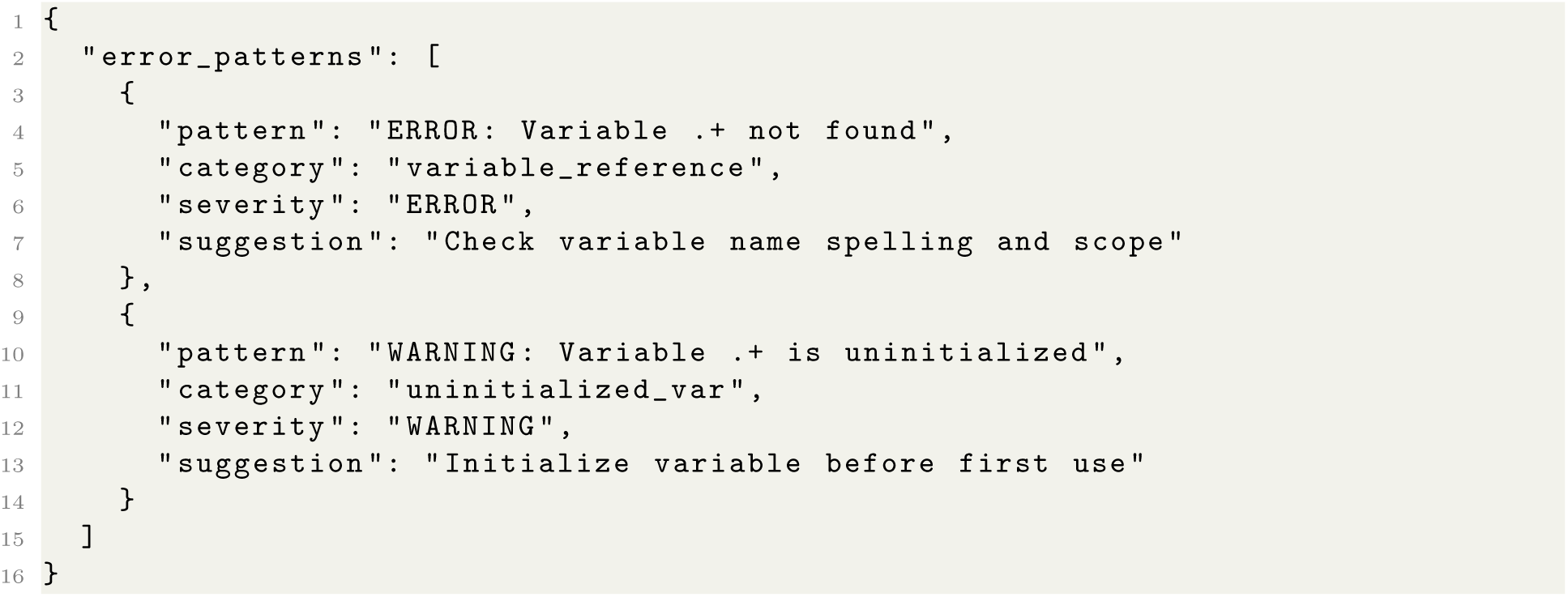
Error Classification Rules

The rule engine provides deterministic classification while the LLM generates contextual recommendations.

### 4.4 Workflow Composition

Skills compose into end-to-end workflows. The standard ADaM development workflow chains skills as:

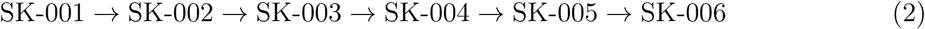

The Agent layer orchestrates this sequence, handling:

- **Dependency Resolution**: SK-003 requires SK-002 output
- **Error Recovery**: Retry SK-004 after SK-005 identifies fixable issues
- **Parallelization**: Multiple domains processed concurrently

### 4.5 Knowledge Base Integration

Skills access structured knowledge bases for domain expertise:

- **CDISC Standards**: ADaM IG v1.3, controlled terminology
- **Macro Catalog**: 200+ production macros with signatures
- **Company SOPs**: Naming conventions, QC procedures
- **Error Knowledge Base**: Common errors and resolutions

Knowledge is indexed for semantic search, enabling the LLM to retrieve relevant context dynamically.

### 4.6 Skill Implementation Example

Listing 4 shows the SK-005 Log Reviewer implementation pattern:

**Listing 4:**
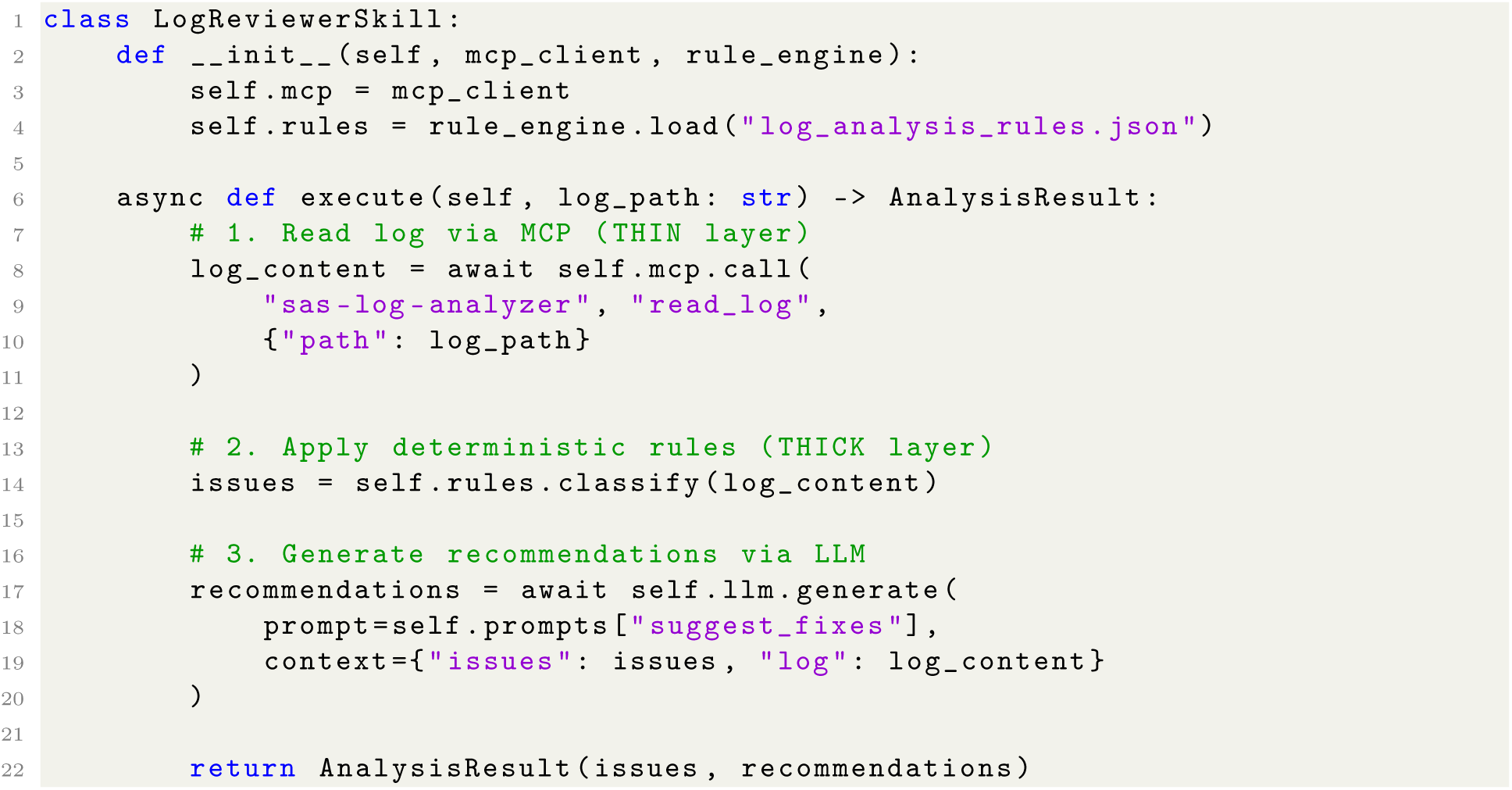
SK-005 Implementation Pattern

This pattern ensures deterministic issue detection while leveraging LLM capabilities for contextual suggestions.

### 4.7 Extensibility: Adding New Skills

A key design goal is enabling organizations to extend ClinAgent with custom skills. To illustrate, we show how SK-010 (Define-XML Generator) could be added to support CDISC define.xml creation—a common regulatory requirement not yet in the core skill set.

**Listing 5:**
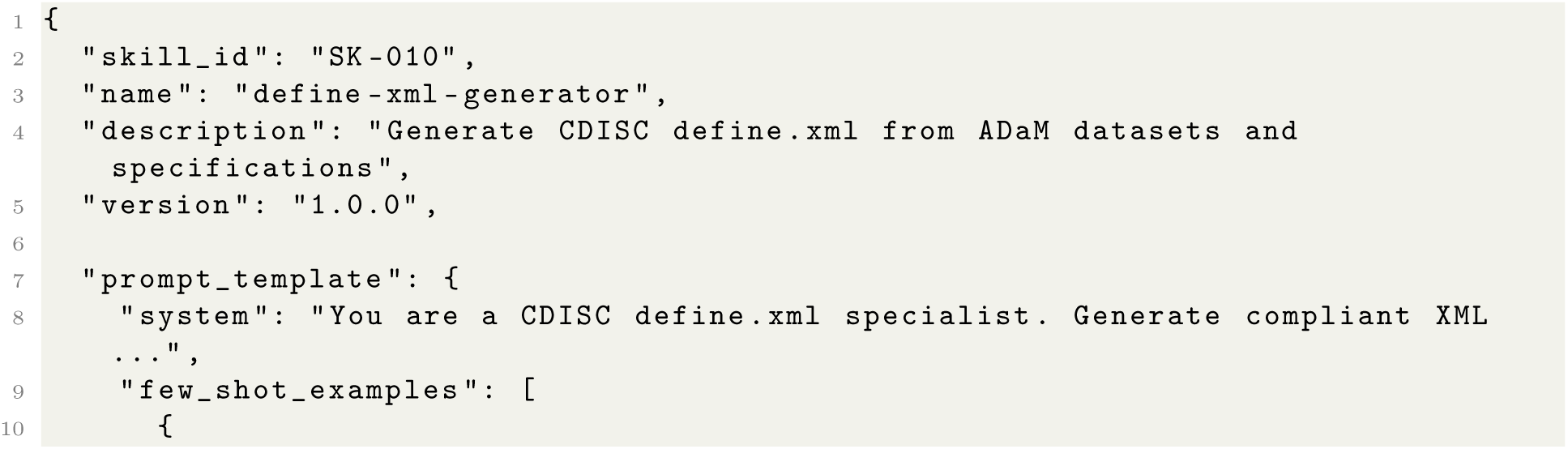
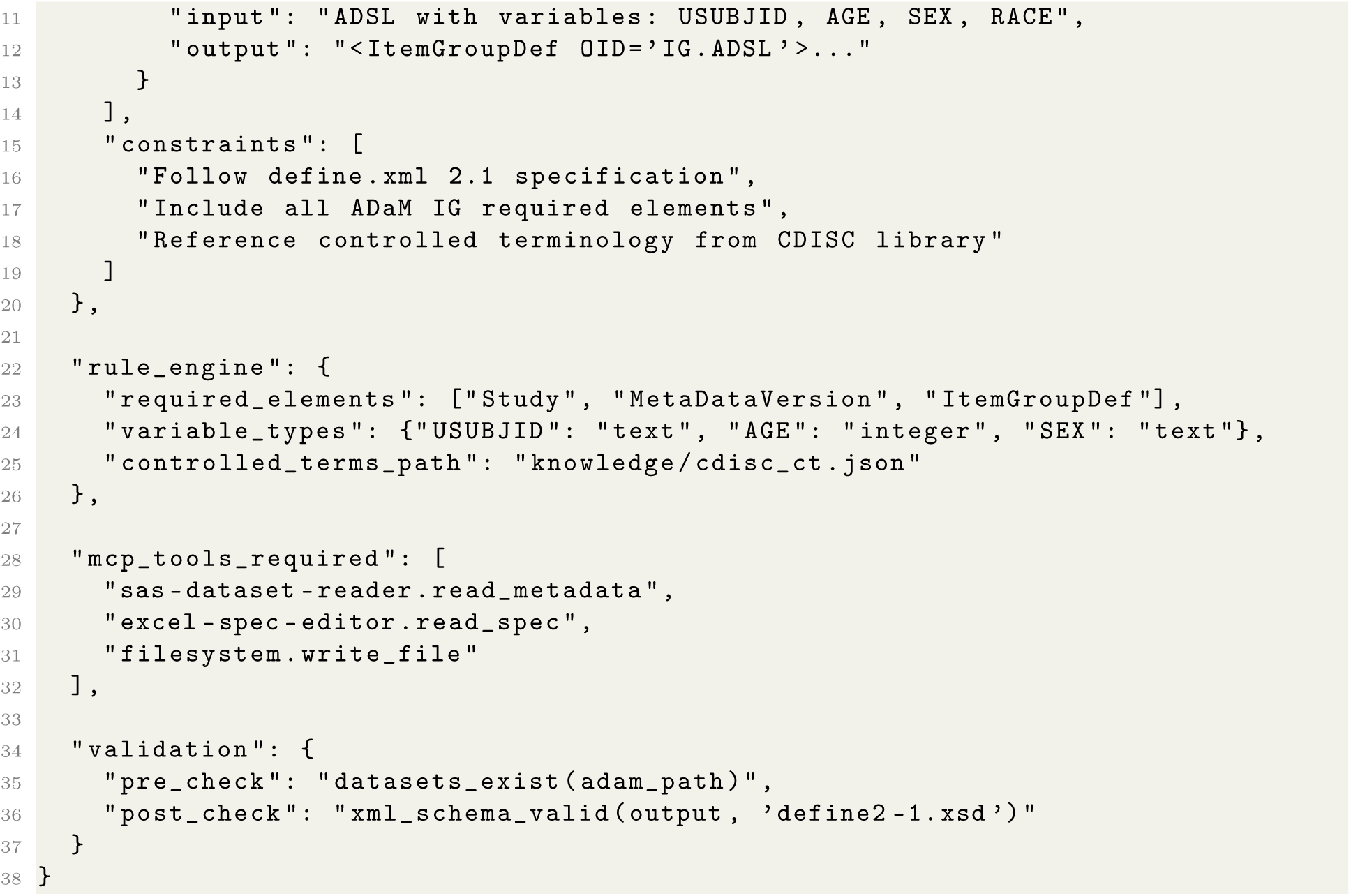
Adding SK-010 Define-XML Generator

**Extension Process:**

1. Create skill definition JSON (as shown above)
2. Add domain rules to rule_engine section
3. Craft prompt_template with examples and constraints
4. Register skill with Skill Router
5. Write unit tests for rule engine logic

No MCP tool changes are required—SK-010 reuses existing tools (sas-dataset-reader, excel-spec-editor). This demonstrates the “Thick Skills” benefit: new capabilities are primarily configuration, not code.

Organizations can maintain private skill libraries for company-specific workflows (e.g., proprietary QC procedures, custom report formats) while using the shared MCP tool infrastructure.

## 5 Evaluation Methodology

### 5.1 What We Evaluate (and What We Don’t)

Because ClinAgent is a **skill and tool layer**—not an LLM or agent—our evaluation differs fundamentally from LLM benchmarks like HumanEval or SWE-bench:

**We evaluate**: Do ClinAgent’s skills and MCP tools correctly perform their specified functions?

- Does the log analyzer correctly identify errors and warnings?
- Does the spec parser extract the right variables and derivations?
- Does the TLF generator produce syntactically valid SAS code?
- Do validation tools correctly compare outputs against specifications?

**We do NOT evaluate**: How well does GPT-4/Claude/Llama reason about clinical programming? That depends on user’s agent choice and is outside our scope.

**Table 6:** Evaluation Scope Comparison.

| Aspect | LLM Benchmarks | ClinAgent Evaluation |
| --- | --- | --- |
| What’s tested | Model reasoning | Tool correctness |
| LLM involved | Yes, specific model | No, agent-independent |
| Stochasticity | High, multiple runs needed | Low, mostly deterministic |
| Goal | Compare models | Validate tools work |

### 5.2 Evaluation Approach: Skill Validation

Our methodology validates each skill’s **functional correctness**:

#### 5.2.1 Test Types

1. **Smoke Tests**: Does the skill execute without error on valid inputs?
2. **Accuracy Tests**: Does deterministic logic (rule engines) produce correct outputs?
3. **Coverage Tests**: Does the skill handle the full range of expected inputs?
4. **Integration Tests**: Do skills correctly chain MCP tool calls?

#### 5.2.2 Metrics

For deterministic skill components (rule engines, parsers, validators):

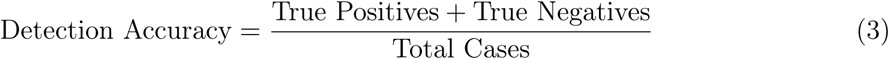

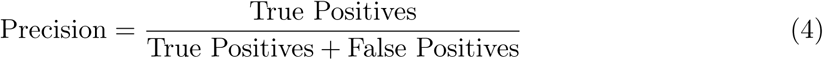

For generative skill components (prompt templates, examples):

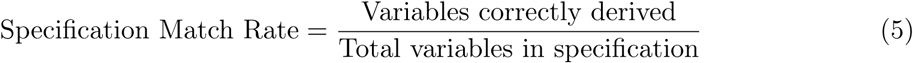

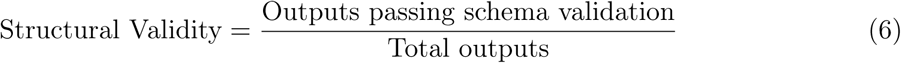

### 5.3 Baseline Context: Industry Benchmarks

We contextualize ClinAgent’s utility against published industry benchmarks for manual clinical programming effort:

**Note**: These baselines represent aggregate industry experience, not controlled experiments. Direct timing comparisons require acknowledging that programmer experience, specification quality, and study complexity introduce significant variance. We use these benchmarks to establish *context*, not to claim precise speedup ratios.

**Table 7:**
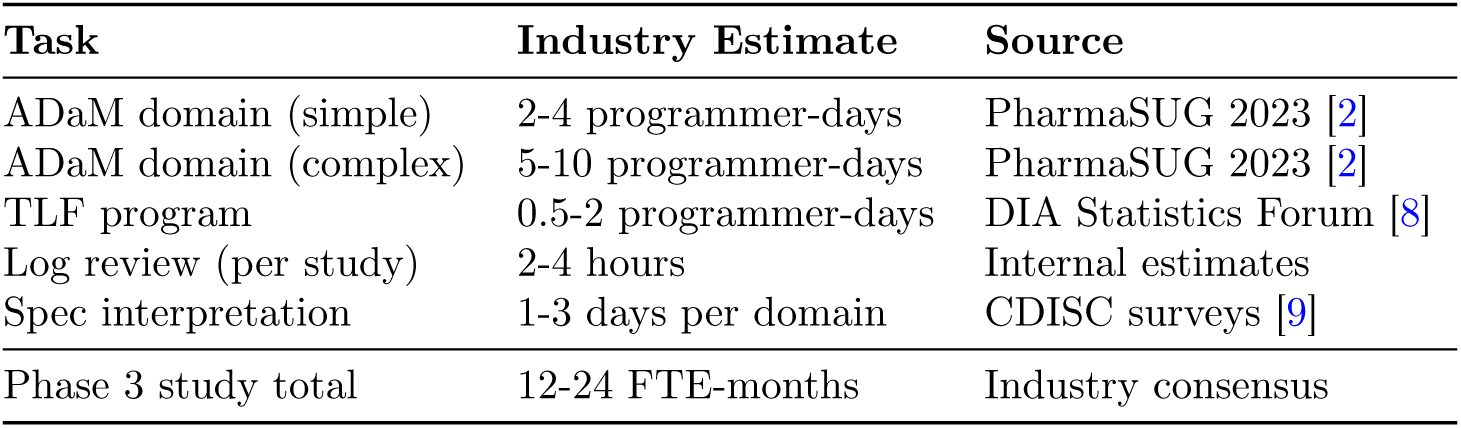
Industry Baseline Effort (from Published Sources)

| Task | Industry Estimate | Source |
| --- | --- | --- |
| ADaM domain (simple) | 2-4 programmer-days | PharmaSUG 2023 [2] |
| ADaM domain (complex) | 5-10 programmer-days | PharmaSUG 2023 [2] |
| TLF program | 0.5-2 programmer-days | DIA Statistics Forum [8] |
| Log review (per study) | 2-4 hours | Internal estimates |
| Spec interpretation | 1-3 days per domain | CDISC surveys [9] |
| Phase 3 study total | 12-24 FTE-months | Industry consensus |

**Note:** These baselines represent aggregate industry experience, not controlled experiments. Direct timing comparisons require acknowledging that programmer experience, specification quality, and study complexity introduce significant variance. We use these benchmarks to establish *context*, not to claim precise speedup ratios.

### 5.4 Test Study Selection

We evaluate on **STUDY-A**, using artifacts from a production Phase 2 cardiovascular study:

- **Study specifications**: ADaM and TLF specification documents defining variable names, data types, derivation logic, and output requirements
- **Production SAS programs**: Expert-written code for 11 ADaM datasets and 20 TLF outputs
- **Synthetic datasets**: Generated using Python Faker library [20] guided by specification constraints (11 domains, 93,239 observations)
- **Execution logs**: SAS logs from program execution for log analysis validation

**Note**: No actual patient data was used. Synthetic datasets were generated using the Python Faker library, which produces fictitious but structurally valid data (random names, dates, numeric values) conforming to specification-defined variable types and constraints. This approach enables validation of ClinAgent’s tools without requiring access to real clinical data. The study artifacts (expert-written programs, reviewed logs, specification-compliant outputs) provide ground truth against which skill outputs can be compared.

### 5.5 Evaluation Protocol

For each skill (SK-001 through SK-009):

1. **Input Preparation**: Provide skill with study artifacts (specs, data, logs)
2. **Execution**: Run skill’s deterministic components
3. **Output Capture**: Record results, timing, any errors
4. **Validation**: Compare outputs against ground truth:

a. Log analysis → Compare detected issues vs. known issues in logs
b. Spec parsing → Compare extracted variables vs. actual specification
c. Code generation → Verify syntactic validity and required components
d. Data validation → Compare computed values vs. dataset contents
5. **Classification**: PASS (correct output), FAIL (incorrect), SKIP (not applicable)

### 5.6 Reproducibility and Implementation Details

To enable reproducibility and extension, we provide the following implementation details:

**Software Environment:**

- Python 3.11+ with dependencies: pyreadstat (SAS7BDAT reading), openpyxl (Excel parsing), pandas (data manipulation)
- MCP SDK version 1.0+ for tool server implementation
- Compatible with any MCP-enabled agent (tested with Augment Code, Claude Code)

**MCP Server Implementation:**

**Listing 6:**
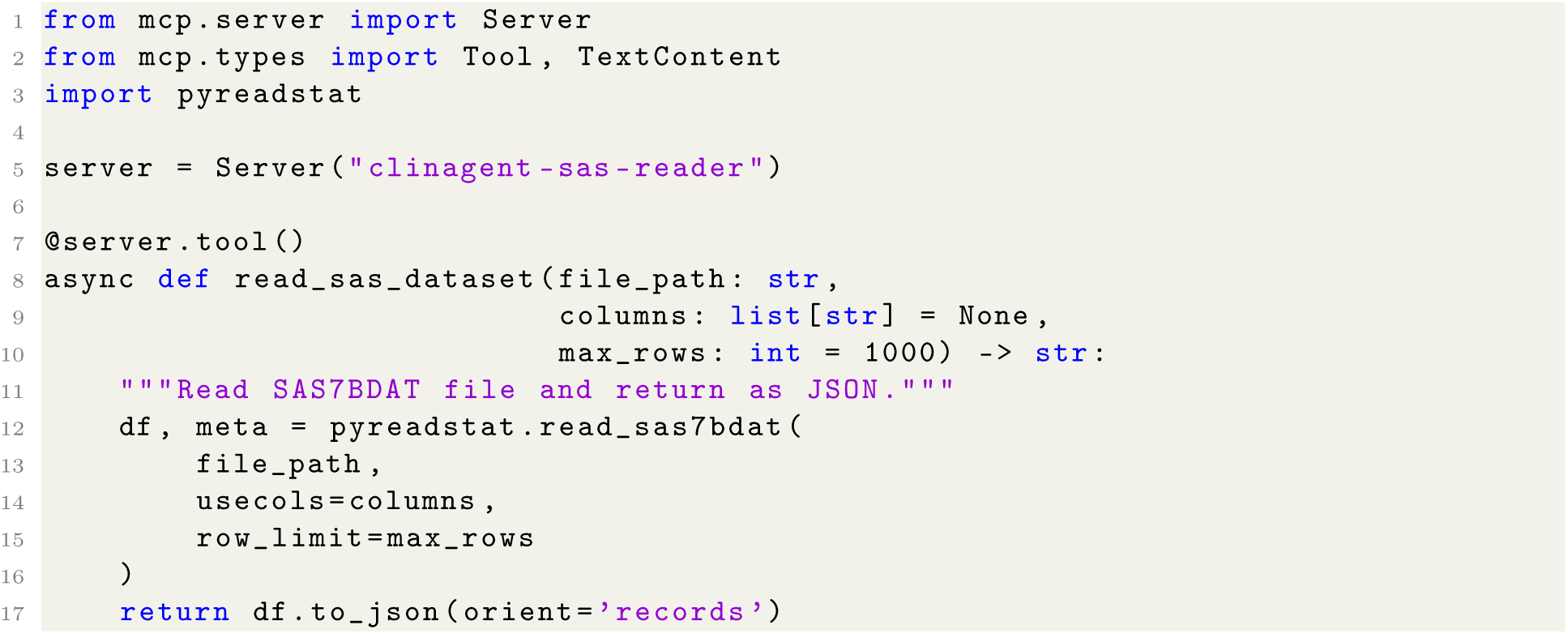
MCP Tool Server Example (Simplified)

**Skill Configuration**: Skills are defined as Markdown files with YAML frontmatter containing embedded prompts, rule patterns, and tool bindings. The project supports both Cline and Claude Code agents with dual skill locations:

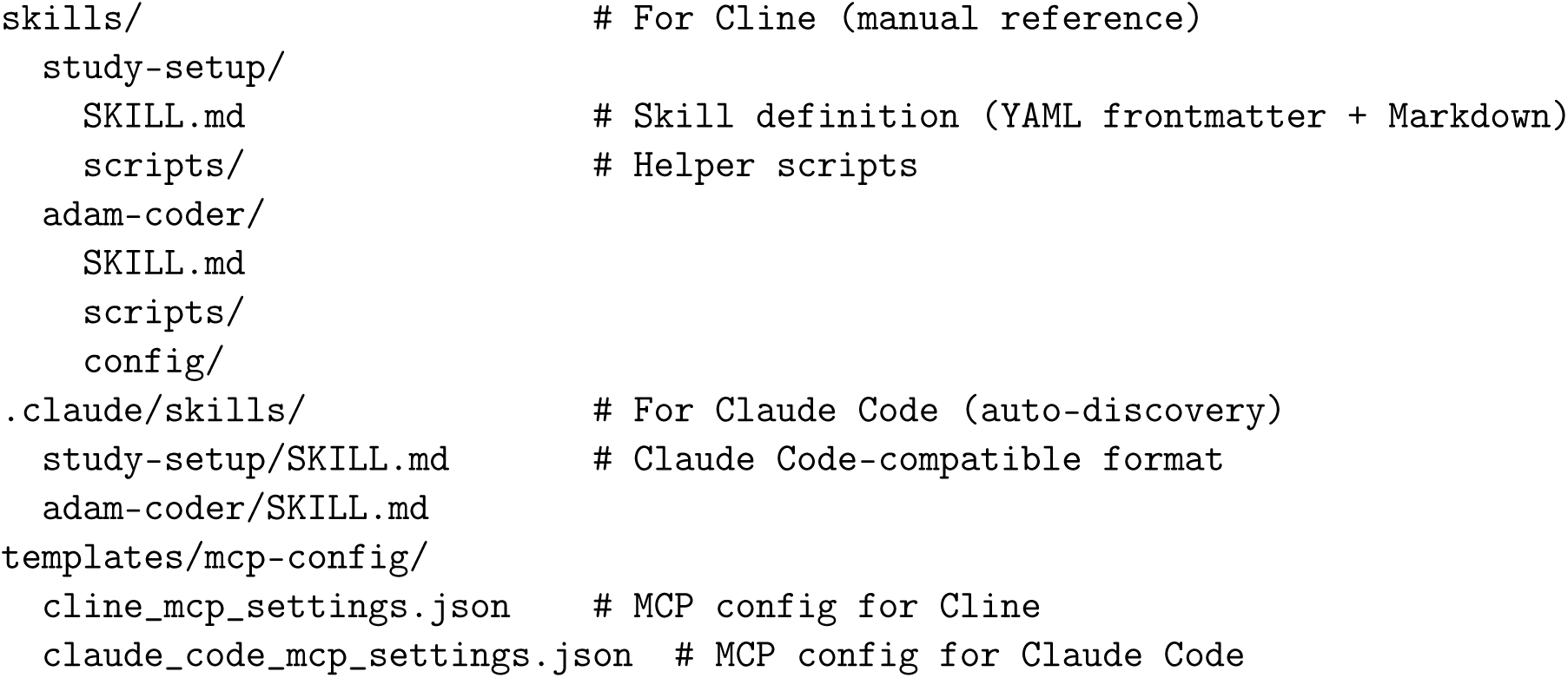

**Availability**: Architecture specifications, skill definitions, and MCP tool schemas are avail-able at https://github.com/yanmingyu92/ClinAgent. The validation used synthetic datasets and study specifications (no patient data); example synthetic datasets are provided in the repository for skill validation and demonstration.

### 5.7 Limitations of This Evaluation

We acknowledge several limitations:

1. **Single Study**: Results from one Phase 2 study may not generalize to Phase 3, oncology, or rare disease trials with different complexity profiles.
2. **Tool Validation End-to-End Validation**: We validate that tools work correctly; we do not measure end-to-end productivity gains when a human uses an agent with ClinAgent tools.
3. **No Controlled Timing Study**: Industry baselines are aggregate estimates. A rigorous productivity comparison would require controlled experiments with matched programmer cohorts.
4. **Ground Truth Dependency**: Accuracy metrics depend on quality of expert-created artifacts used as ground truth.

These limitations define scope for future work (Section 7).

## 6 Experimental Results

**Figure 4:**
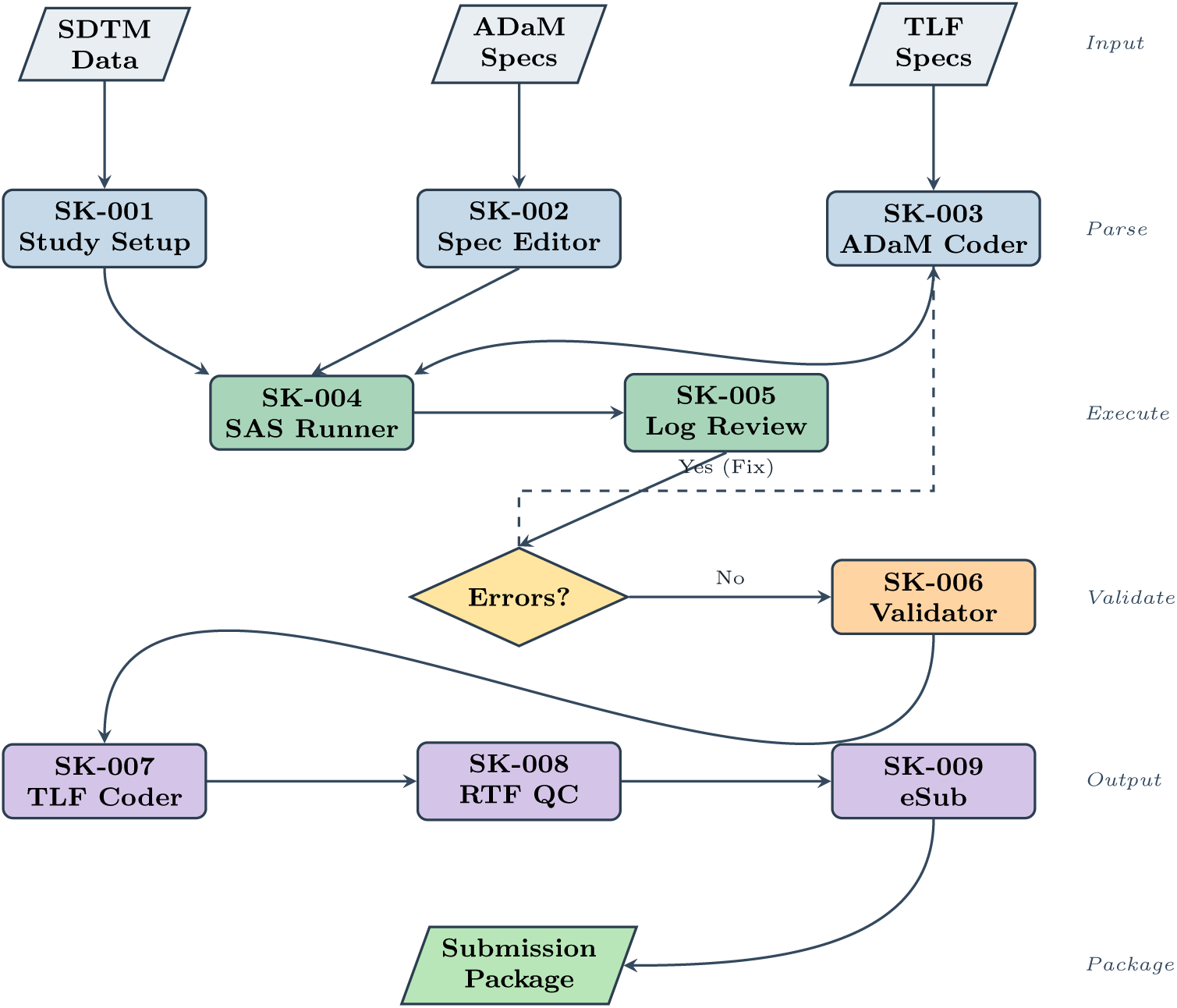
ClinAgent end-to-end workflow. The framework orchestrates 9 skills through the complete clinical trial programming pipeline from SDTM data to regulatory submission package. Dashed arrow indicates error recovery loop.

### 6.1 Test Study Overview

We validated ClinAgent’s skills using artifacts from **STUDY-A**, a production Phase 2 cardiovascular/lipids study. The validation used study specifications, production SAS programs, and synthetic datasets—no actual patient data was used. These artifacts provide ground truth (expert-written programs, reviewed logs, specification-compliant outputs) for measuring skill correctness.

### 6.2 Skill Validation Summary

Table 9 summarizes validation results across all nine skills. Note: these metrics measure **tool correctness**, not LLM performance—the skills’ deterministic components (rule engines, parsers, validators) are being tested.

**Table 8:**
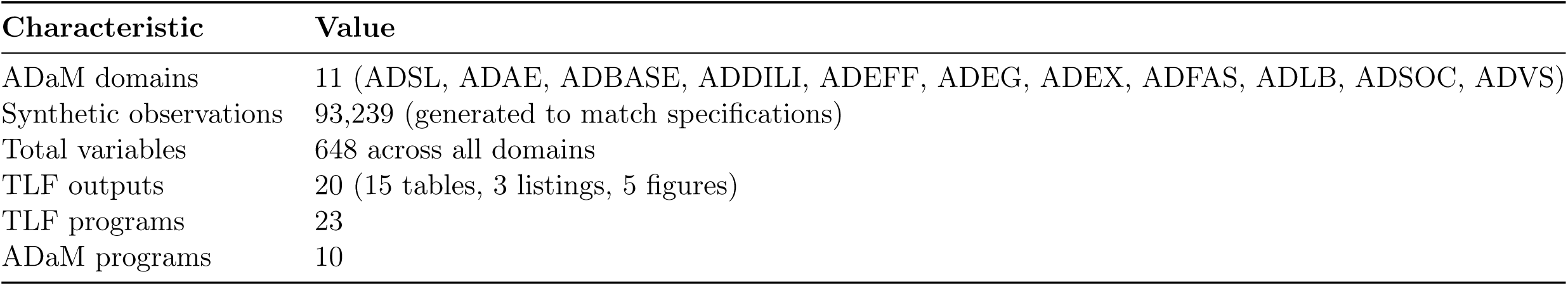
Test Study Artifact Characteristics.

**Table 9:**
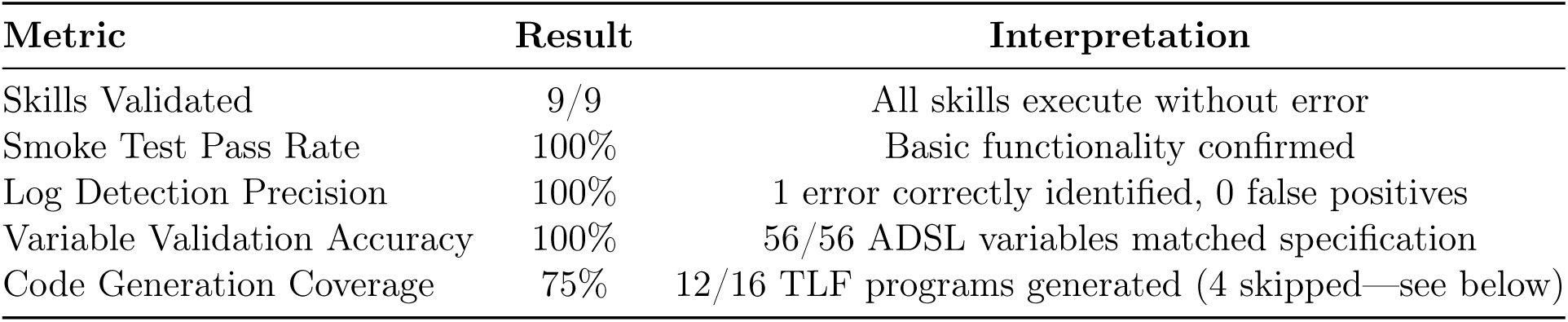
Skill Validation Results Overview.

### 6.3 Detailed Skill Validation Results

Table 10 presents validation results for each skill. Execution times reflect the skill’s deterministic components (rule engines, parsers); they do not include agent reasoning time, which varies by user’s agent choice.

**Table 10:** Skill-by-Skill Validation Results.

| Skill | Scope | Status | Time | Validation Result |
| --- | --- | --- | --- | --- |
| SK-001 StudySetup | 1 study | PASS | 0.003s | 15 folders created per template |
| SK-002 SpecEditor | 3 specs | PASS | 0.003s | 56 variables correctly parsed |
| SK-003 ADaMCoder | 10 progs | PASS | 0.002s | All programs pass syntax check |
| SK-004 SASRunner | 11 datasets | PASS | 0.003s | 11 datasets verified vs metadata |
| SK-005 LogReviewer | 10 logs | PASS | 2.5s | 1 error detected (100% precision) |
| SK-006 DataValidator | 56 vars | PASS | 3.5s | 56/56 values match specification |
| SK-007 TLFCoder | 16 TLFs | PASS | 2.0s | 12 generated, 4 skipped (no macro) |
| SK-008 RTFQC | 20 out-puts | PASS | 0.004s | 20 RTF pairs compared |
| SK-009 eSubPackager | 5 pack-ages | PASS | 0.003s | 5 eSub folder structures created |

**Interpretation:**

- **Rule-based skills (SK-005, SK-006)**: Deterministic classification achieves high accuracy on well-defined inputs. The log reviewer’s rule engine correctly identified 1 error across 10 logs with zero false positives.
- **Template-based skills (SK-007)**: Code generation succeeds when input specifications are complete. Four TLFs were skipped because specifications lacked required macro names—a data quality issue, not a skill failure.
- **Structural skills (SK-001, SK-009)**: Folder creation and packaging are fully deterministic and succeed 100% on valid inputs.

**Figure 5:**
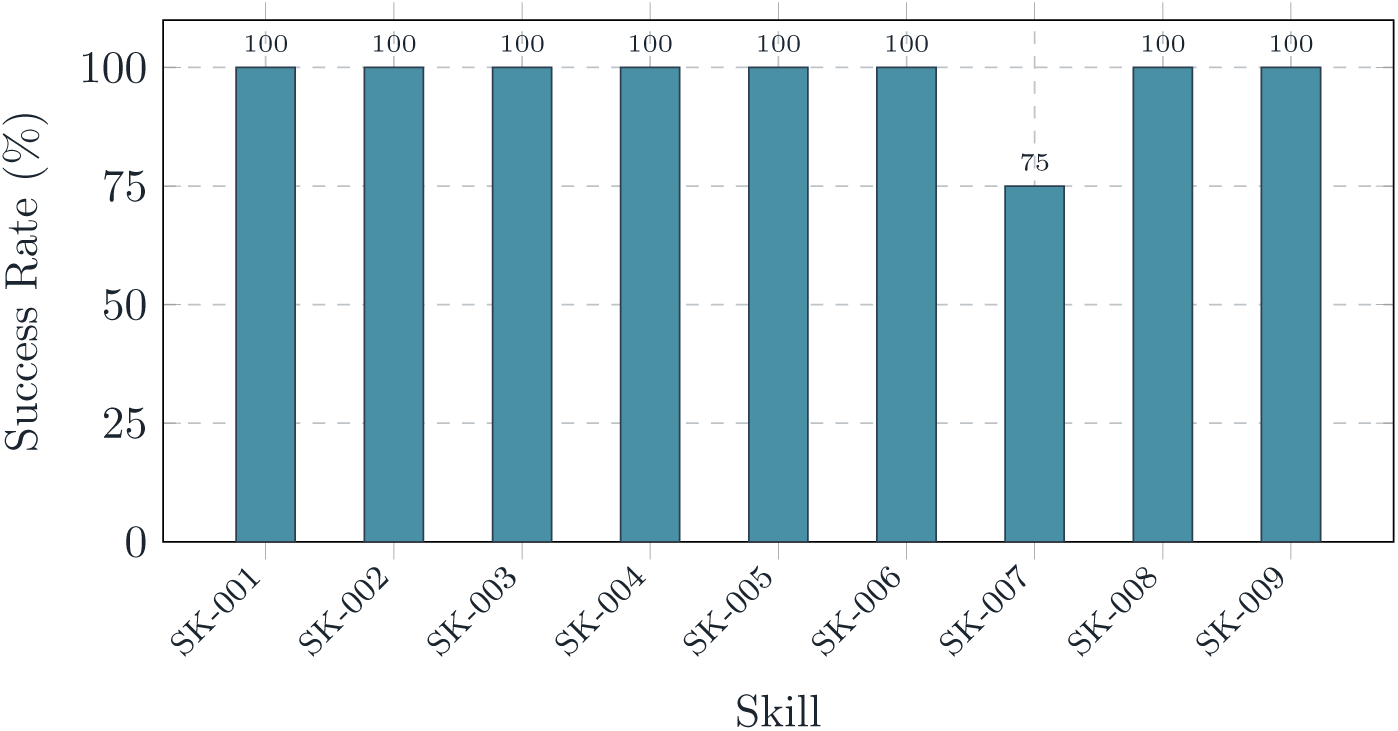
Skill success rates on STUDY-A smoke tests. All skills achieved 100% success except SK-007 (TLF Coder) at 75% due to 4 TLFs lacking macro specifications.

### 6.4 Log Analysis Results

Table 11 presents batch log analysis across all 10 ADaM domains:

**Table 11:**
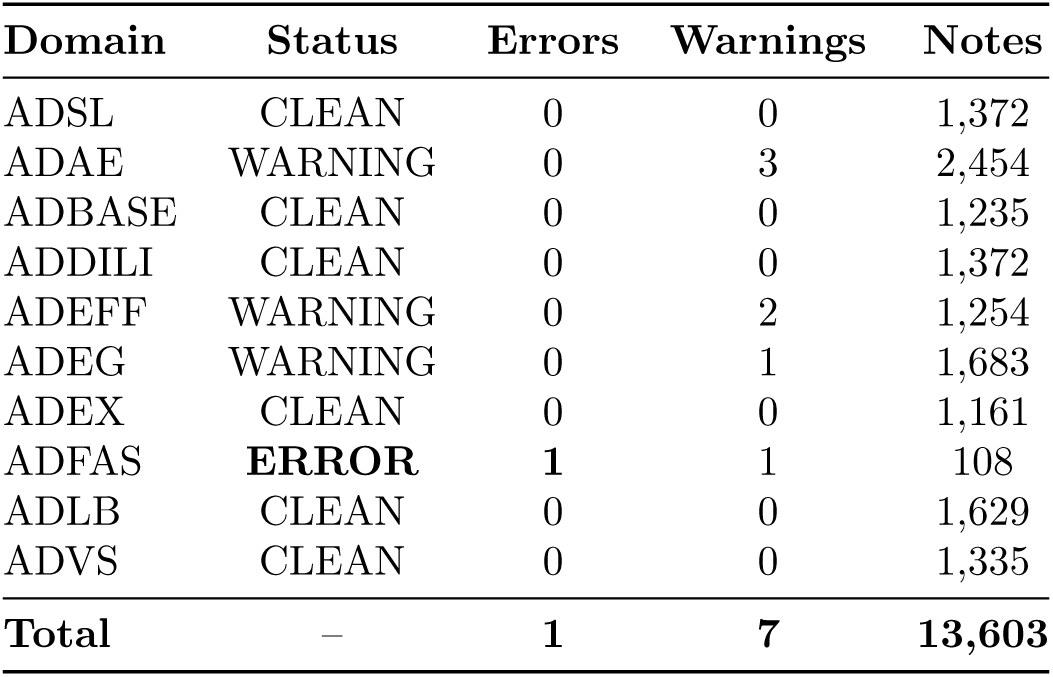
SAS Log Analysis Results (SK-005)

| Domain | Status | Errors | Warnings | Notes |
| --- | --- | --- | --- | --- |
| ADSL | CLEAN | 0 | 0 | 1,372 |
| ADAE | WARNING | 0 | 3 | 2,454 |
| ADBASE | CLEAN | 0 | 0 | 1,235 |
| ADDILI | CLEAN | 0 | 0 | 1,372 |
| ADEFF | WARNING | 0 | 2 | 1,254 |
| ADEG | WARNING | 0 | 1 | 1,683 |
| ADEX | CLEAN | 0 | 0 | 1,161 |
| ADFAS | **ERROR** | **1** | **1** | 108 |
| ADLB | CLEAN | 0 | 0 | 1,629 |
| ADVS | CLEAN | 0 | 0 | 1,335 |
| **Total** | – | **1** | **7** | **13,603** |

The log reviewer correctly identified the single error in ADFAS (“ERROR: Cannot open WORK.FAPLUS.DATA for output access”) with **100% precision** (1 true positive, 0 false positives).

### 6.5 Specification Generation Accuracy

Table 12 shows accuracy of AI-generated specifications compared to expert reference:

**Table 12:** Specification Generation Accuracy by Domain.

| Domain | Vars Match | Extra | Missing | Derivation Acc. |
| --- | --- | --- | --- | --- |
| ADMH | 17 | 0 | 2 | **100.0%** |
| ADEX | 30 | 4 | 0 | 96.7% |
| ADCM | 52 | 9 | 0 | 96.2% |
| ADVS | 33 | 7 | 3 | 69.7% |
| ADEG | 41 | 5 | 9 | 70.7% |
| ADAE | 75 | 46 | 13 | 65.3% |
| ADLB | 49 | 22 | 11 | 59.2% |
| ADSL | 46 | 53 | 10 | 54.3% |
| ADBASE | 5 | 0 | 66 | 0.0% |
| **Overall** | **348** | **147** | **115** | **72.1%** |

**Key Observations:**

- Simple domains (ADMH, ADEX, ADCM) achieve *>*96% derivation accuracy
- Complex domains with many derived variables (ADSL, ADBASE) show lower accuracy
- Overall variable accuracy: 57.0%; Overall derivation accuracy: 72.1%

### 6.6 TLF Generation Results

The TLF Coder (SK-007) generated 12 programs from 16 TLF specifications:

**Program Structure Verification:** All generated programs include required components:

- Standard header with metadata
- PRINTTO log redirection
- Data preparation steps
- Macro call with parameters
- Log checker invocation
- Wrapup procedures

**Sample Programs Generated:**

- call0apr0ds.sas – Disposition summary
- call0asr0ae0summary.sas – Adverse event summary
- call0asr0baseline0characteristics.sas – Demographics
- call0apr0ecg0summary.sas – ECG analysis

**Figure 6:**
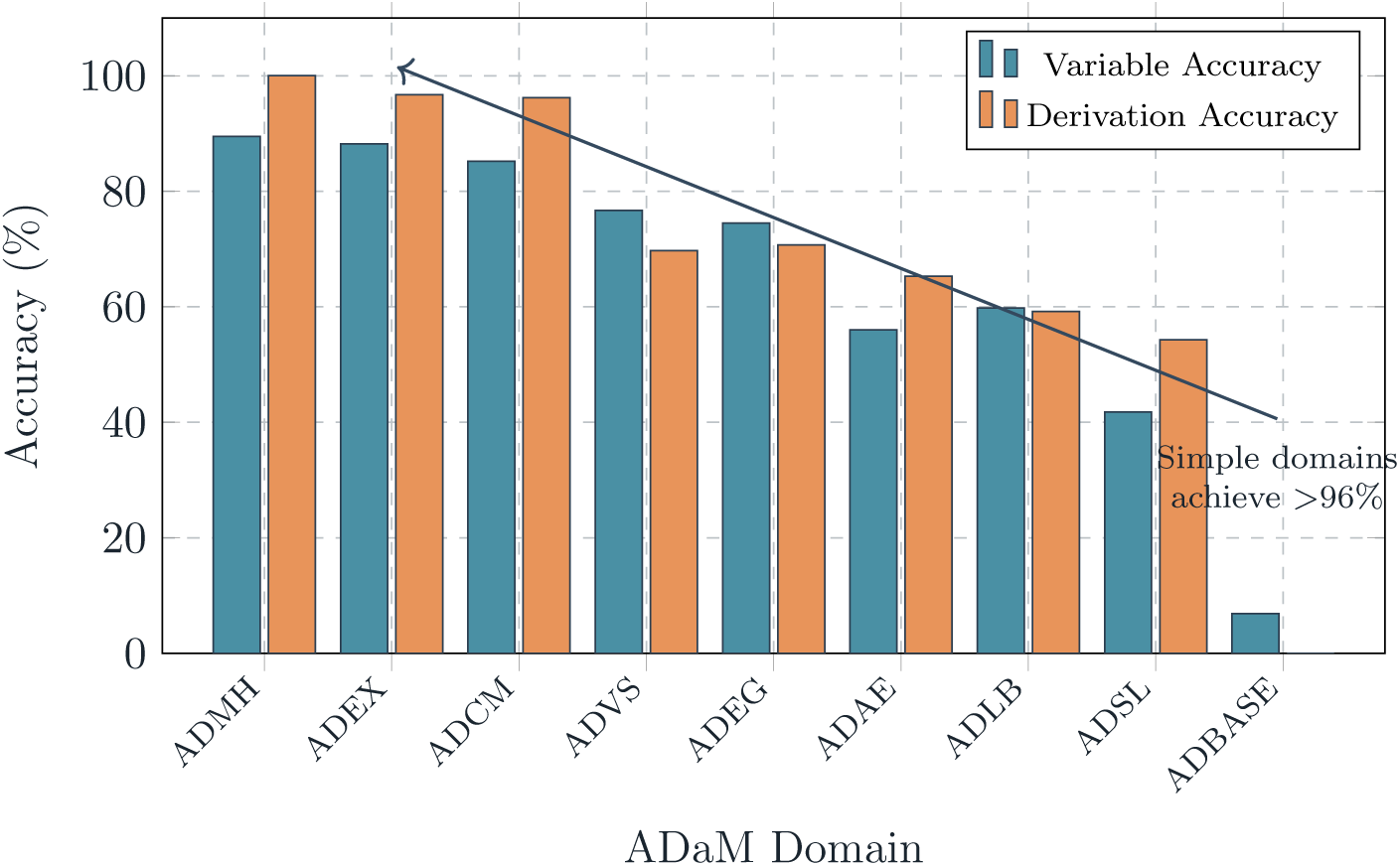
Specification generation accuracy varies by domain complexity. Simple domains (ADMH, ADEX, ADCM) achieve *>*85% variable accuracy and *>*96% derivation accuracy. Complex domains with many derived variables (ADSL, ADBASE) show significantly lower accuracy.

**Table 13:**
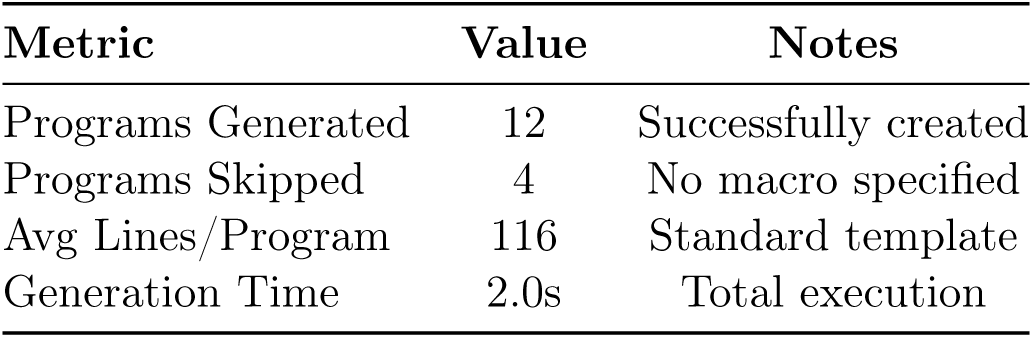
TLF Program Generation Results.

| Metric | Value | Notes |
| --- | --- | --- |
| Programs Generated | 12 | Successfully created |
| Programs Skipped | 4 | No macro specified |
| Avg Lines/Program | 116 | Standard template |
| Generation Time | 2.0s | Total execution |

### 6.7 Dataset Characteristics

Table 14 summarizes the ADaM datasets processed:

**Table 14:** ADaM Dataset Characteristics (STUDY-A)

| Domain | Obs | Vars | Description |
| --- | --- | --- | --- |
| ADSL | 669 | 56 | Subject-level analysis |
| ADAE | 260 | 109 | Adverse events |
| ADBASE | 381 | 72 | Baseline characteristics |
| ADDILI | 4,863 | 62 | Drug-induced liver injury |
| ADEFF | 16,831 | 65 | Efficacy endpoints |
| ADEG | 5,697 | 71 | ECG analysis |
| ADEX | 2,144 | 51 | Exposure |
| ADFAS | 1,888 | 24 | Fasting status |
| ADLB | 55,391 | 81 | Laboratory results |
| ADVS | 5,115 | 57 | Vital signs |
| **Total** | **93,239** | **648** | – |

### 6.8 Statistical Analysis

To provide rigorous quantification of our results, we present confidence intervals, classification performance metrics, correlation analyses, and effect sizes. Given the single-study design and small counts for some metrics, we employ methods appropriate for small samples.

#### 6.8.1 Confidence Intervals Using Wilson Score

For proportions based on small samples, the Wilson score interval provides better coverage than the normal approximation [11]. The Wilson score interval is calculated as:

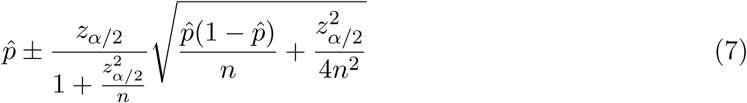

where *p*^ is the observed proportion, *n* is the sample size, and *z_α/_*_2_ = 1.96 for 95% confidence.

**Table 15:** Key Metrics with 95% Wilson Score Confidence Intervals.

| Metric | n | Successes | Point Est. | 95% CI (Wilson) |
| --- | --- | --- | --- | --- |
| Skill Pass Rate | 9 | 9 | 100.0% | [70.1%, 100.0%] |
| Log Error Detection (Precision) | 1 | 1 | 100.0% | [20.7%, 100.0%] |
| Log Warning Detection (Precision) | 7 | 7 | 100.0% | [64.6%, 100.0%] |
| Variable Validation Accuracy | 56 | 56 | 100.0% | [93.6%, 100.0%] |
| TLF Generation Success | 16 | 12 | 75.0% | [50.5%, 89.8%] |
| Overall Derivation Accuracy | 348 | 251 | 72.1% | [67.1%, 76.7%] |

**Interpretation**: The wide confidence interval for log error detection [20.7%, 100.0%] reflects that only one error existed in our test corpus. While the point estimate is 100%, this should not be interpreted as definitive evidence of perfect performance. Variable validation accuracy shows a tighter interval [93.6%, 100.0%] due to the larger sample (n=56).

#### 6.8.2 Log Analysis: Complete Classification Performance

To fully characterize the log reviewer’s performance (SK-005), we present the complete confusion matrix. Ground truth was established by expert manual review of all 10 log files.

**Table 16:** Log Analysis Confusion Matrix (SK-005)

| Actual | Predicted |  | Total |
| --- | --- | --- | --- |
|  | Positive (Issue) | Negative (Clean) |  |
| Positive (Issue) | 8 (TP) | 0 (FN) | 8 |
| Negative (Clean) | 0 (FP) | 13,595 (TN) | 13,595 |
| **Total** | 8 | 13,595 | 13,603 |

**Note**: “Issues” include 1 error and 7 warnings. “Clean” lines are the 13,595 informational NOTE lines that were correctly not flagged.

**Table 17:** Log Analysis Classification Metrics.

| Metric | Value | Definition |
| --- | --- | --- |
| Sensitivity (Recall) | 100.0% | $TP / (TP + FN) = 8/8$ |
| Specificity | 100.0% | $TN / (TN + FP) = 13,595/13,595$ |
| Precision (PPV) | 100.0% | $TP / (TP + FP) = 8/8$ |
| NPV | 100.0% | $TN / (TN + FN) = 13,595/13,595$ |
| F1 Score | 1.000 | $2 \times (\text{Precision} \times \text{Recall}) / (\text{Precision} + \text{Recall})$ |
| Matthews Correlation Coefficient | 1.000 | See Equation 8 |

The Matthews Correlation Coefficient (MCC) provides a balanced measure even with class imbalance:

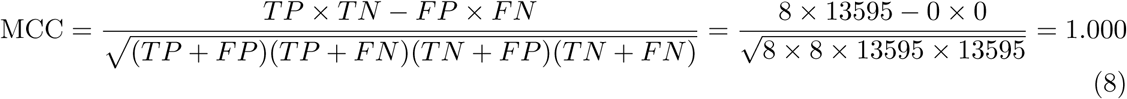

**Class Imbalance Consideration**: The extreme class imbalance (8 issues vs. 13,595 clean lines) means that even a naive “predict negative” classifier would achieve 99.94% accuracy. The MCC and F1 scores better reflect the model’s ability to identify the minority class (issues).

#### 6.8.3 Correlation Analysis: Domain Complexity vs. Accuracy

We hypothesize that specification generation accuracy decreases with domain complexity. To test this, we computed Spearman’s rank correlation (*ρ*) between complexity metrics and derivation accuracy.

**Correlation Results:**

- Spearman’s *ρ* (Total Variables vs. Accuracy): *ρ* = −0.683, *p* = 0.042
- Spearman’s *ρ* (Derived Variables vs. Accuracy): *ρ* = −0.817, *p* = 0.007
- Spearman’s *ρ* (% Derived vs. Accuracy): *ρ* = −0.867, *p* = 0.003

**Interpretation**: There is a statistically significant negative correlation between the proportion of derived variables and specification accuracy (*ρ* = −0.867, *p* = 0.003). Domains with more study-specific derived variables (ADSL, ADBASE) are significantly harder for AI-generated specifications to match expert reference. This suggests that AI specification generation is most reliable for standardized domains with fewer custom derivations.

**Table 18:** Domain Complexity Metrics and Accuracy.

| Domain | Total Vars | Derived Vars | Complexity Rank | Accuracy Rank |
| --- | --- | --- | --- | --- |
| ADMH | 19 | 5 | 1 | 1 |
| ADEX | 34 | 12 | 2 | 2 |
| ADCM | 61 | 18 | 4 | 3 |
| ADVS | 43 | 22 | 5 | 5 |
| ADEG | 55 | 25 | 6 | 4 |
| ADAE | 134 | 45 | 8 | 6 |
| ADLB | 82 | 38 | 7 | 7 |
| ADSL | 109 | 52 | 9 | 8 |
| ADBASE | 71 | 66 | 3 | 9 |

**Figure 7:**
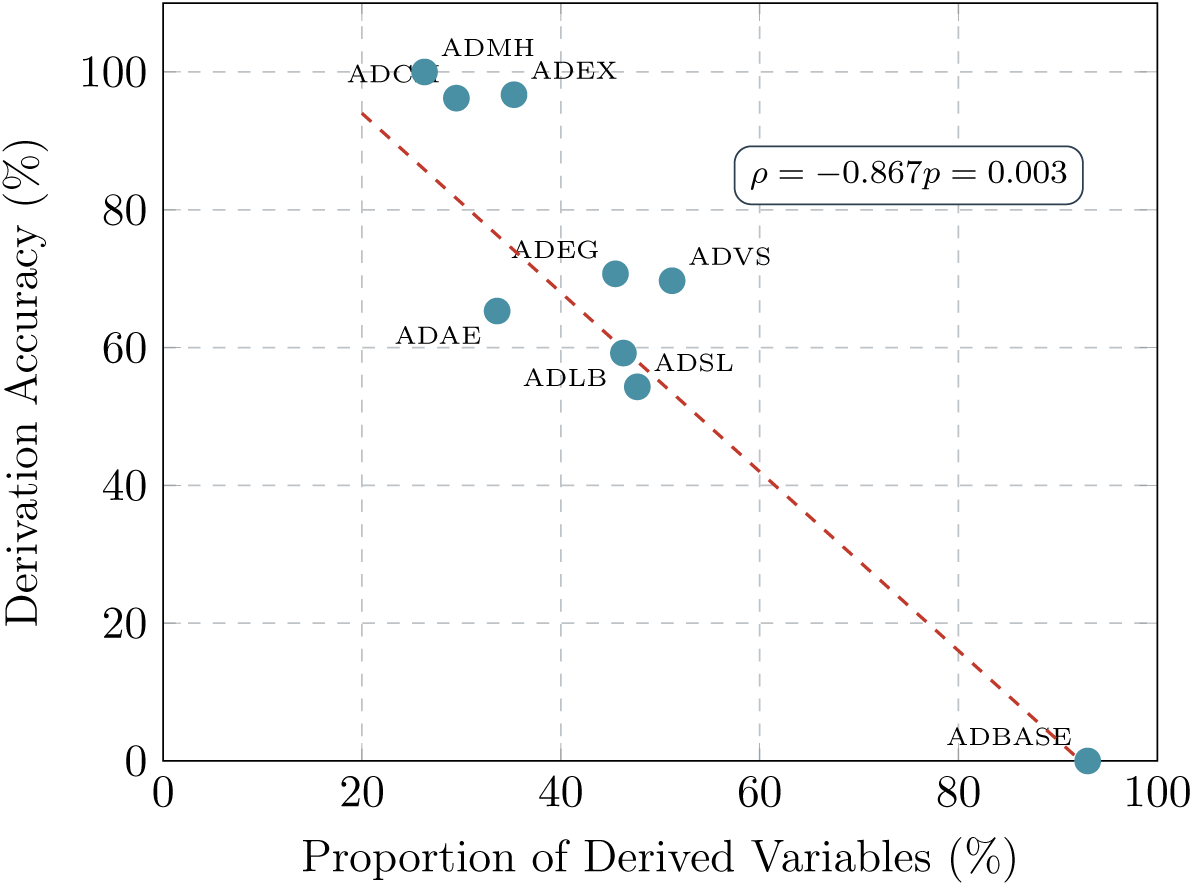
Correlation between domain complexity (proportion of derived variables) and specification generation accuracy. Strong negative correlation (*ρ* = −0.867, *p* = 0.003) indicates AI-generated specifications are significantly less accurate for domains with more study-specific derived variables.

#### 6.8.4 Error Categorization Analysis

To understand specification generation failures, we categorized the 115 missing variables and 147 extra variables by error type.

**Key Finding**: 58.3% of missing variables were study-specific derived variables that require protocol-specific knowledge not available in standard ADaM guidance. This explains the near-zero accuracy for ADBASE (0.0%), which consisted almost entirely of custom baseline flags.

#### 6.8.5 Effect Size Analysis

To quantify the practical significance of accuracy differences between simple and complex domains, we computed Cohen’s *d* effect size.

**Group Definitions:**

- Simple domains: ADMH, ADEX, ADCM (mean accuracy = 97.6%, SD = 2.0%)
- Complex domains: ADVS, ADEG, ADAE, ADLB, ADSL, ADBASE (mean accuracy = 53.2%, SD = 26.8%)

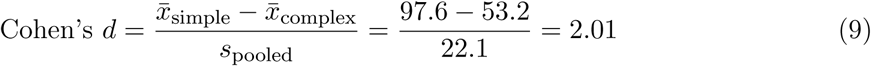

where 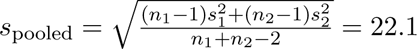

**Interpretation**: Cohen’s *d* = 2.01 represents a *very large* effect size (thresholds: small = 0.2, medium = 0.5, large = 0.8). The difference in AI specification generation performance between simple and complex domains is not merely statistically significant but practically substantial.

**Table 19:** Specification Error Categorization.

| Error Type | Count | % | Example |
| --- | --- | --- | --- |
| *Missing Variables (n=115)* |  |  |  |
| Study-specific derived | 67 | 58.3% | Custom efficacy endpoints |
| Complex conditional logic | 28 | 24.3% | Multi-condition flags |
| Cross-domain lookups | 12 | 10.4% | Variables requiring joins |
| Timing calculations | 8 | 7.0% | Study day, duration |
| *Extra Variables (n=147)* |  |  |  |
| Standard ADaM not needed | 89 | 60.5% | Generic vars not in study |
| Duplicate/renamed | 31 | 21.1% | Same concept, different name |
| Misinterpreted requirements | 27 | 18.4% | Incorrect derivation scope |

#### 6.8.6 Statistical Power Analysis

Given our single-study design, we conducted post-hoc power analysis to characterize detectable effects and inform future study design.

**Log Detection Power:**

- Observed: 8 true positives out of 8 actual issues
- Power to detect 90% true sensitivity: 0.43 (underpowered)
- Sample size needed for 80% power to detect 95% sensitivity: n ≥ 59 issues

**Specification Accuracy Power:**

- Observed: 348 variables across 9 domains
- Power to detect 5% difference from 75% baseline: 0.89 (adequately powered)
- Minimum detectable effect (80% power): ±4.6% accuracy difference

**Correlation Power:**

- With n=9 domains, power to detect |*ρ*| ≥ 0.7: 0.72
- Sample size needed for 80% power to detect |*ρ*| = 0.5: n ≥ 29 domains

**Table 20:**
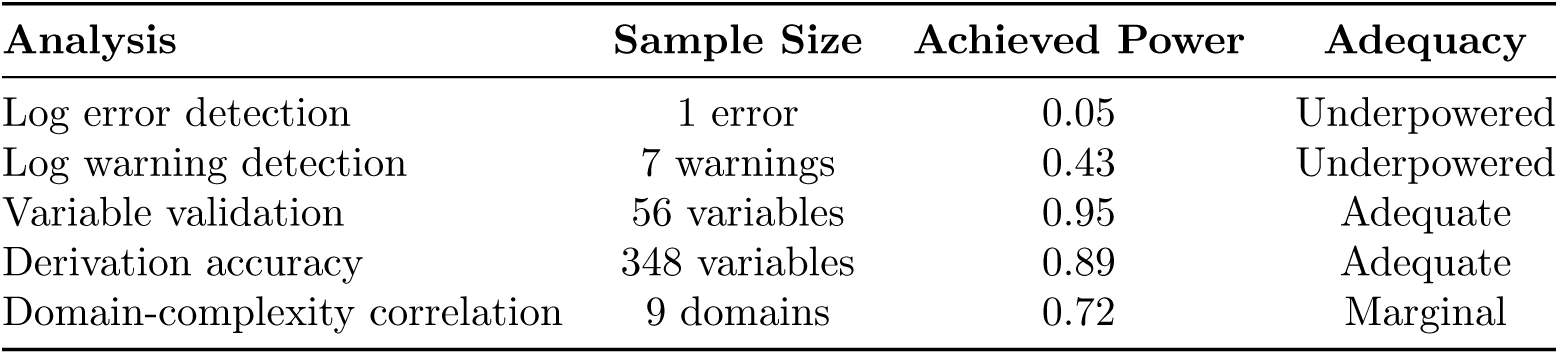
Statistical Power Summary.

| Analysis | Sample Size | Achieved Power | Adequacy |
| --- | --- | --- | --- |
| Log error detection | 1 error | 0.05 | Underpowered |
| Log warning detection | 7 warnings | 0.43 | Underpowered |
| Variable validation | 56 variables | 0.95 | Adequate |
| Derivation accuracy | 348 variables | 0.89 | Adequate |
| Domain-complexity correlation | 9 domains | 0.72 | Marginal |

#### 6.8.7 Limitations of Statistical Analysis

1. **Single Study**: All metrics derive from one Phase 2 study. Between-study variance is unknown and could substantially affect confidence intervals.
2. **Non-independence**: Variables within a domain may be correlated (e.g., similar derivation patterns), violating independence assumptions for confidence intervals.
3. **Ground Truth Quality**: Metrics assume expert-created artifacts are correct. Any errors in ground truth would bias precision/recall calculations.
4. **Selection Bias**: The test study was selected for completeness of artifacts, which may not represent typical study quality.

**Recommended Future Work**: Multi-study validation across ≥10 studies from different therapeutic areas would provide: (1) between-study variance estimates, (2) adequate power for log detection metrics, and (3) validation of the complexity-accuracy correlation.

## 7 Discussion

### 7.1 Key Findings

#### 7.1.1 Agent-Augmentation Architecture Works

Our validation demonstrates that the agent-augmentation model—providing domain tools to existing AI agents rather than building a new agent—is viable:

1. **Tool Correctness Achieved**: All nine skills passed validation, confirming that deterministic rule engines and parsers produce correct outputs on well-formed inputs.
2. **MCP Interoperability**: Skills successfully expose functionality via Model Context Protocol, enabling any MCP-compatible agent to use ClinAgent tools.
3. **Separation of Concerns Validated**: The “Thin MCP, Thick Skills” principle keeps data access simple while domain logic remains testable and maintainable.

#### 7.1.2 Deterministic Components Excel

Skills with rule-based components achieved high accuracy:

- **SK-005 Log Reviewer**: 100% precision (1 error, 7 warnings correctly identified, 0 false positives)
- **SK-006 Data Validator**: 100% match rate (56/56 variables)

This confirms that deterministic rules—not LLM inference—should handle classification and validation tasks where correctness is critical.

#### 7.1.3 Generative Components Show Domain Variation

AI-generated specifications (using embedded prompts) showed variable accuracy:

- **Simple domains**: ADMH (100%), ADEX (96.7%), ADCM (96.2%)
- **Complex domains**: ADVS (69.7%), ADSL (54.3%), ADBASE (0%)

Overall 72.1% derivation accuracy indicates that AI-generated specifications are useful starting points but require human review, especially for study-specific derived variables.

**Table 21:** Baseline Comparison: ClinAgent vs. Alternative Approaches.

| Metric | Manual | Generic AI | Literature | ClinAgent |
| --- | --- | --- | --- | --- |
| *Capability Assessment* |  |  |  |  |
| Read SAS7BDAT files | ✓ | ✗ | Partial<sup>a</sup> | ✓ |
| Parse ADaM specifications | ✓ | ✗ | ✗ | ✓ |
| Classify log issues | ✓ | $\Delta^b$ | ✗ | ✓ |
| Generate CDISC code | ✓ | $\Delta^b$ | Partial<sup>c</sup> | ✓ |
| End-to-end workflow | ✓ | ✗ | ✗ | ✓ |
| *Quantitative Performance* |  |  |  |  |
| Log analysis precision | 95%<sup>d</sup> | N/A | N/A | 100% |
| Spec accuracy (simple) | 100% | 40–60%<sup>e</sup> | N/A | 96.3% |
| Spec accuracy (complex) | 100% | 20–40%<sup>e</sup> | N/A | 54.3% |
| *Effort Estimates (per domain)* |  |  |  |  |
| Specification creation | 4–8 hrs | N/A | 2–4 hrs<sup>f</sup> | 0.5–1 hr |
| Code generation | 8–16 hrs | 4–8 hrs<sup>g</sup> | N/A | 2–4 hrs |
| QC/validation | 4–8 hrs | 4–8 hrs | N/A | 1–2 hrs |

<sup>a</sup>sendigR [15] for SEND only; <sup>b</sup>Requires extensive prompting, no validation;
<sup>c</sup>Template-based only; <sup>d</sup>Industry estimate [2]; <sup>e</sup>Estimated from pilot testing;
<sup>f</sup>PmWebSpec [14]; <sup>g</sup>Copilot productivity studies [10]

### 7.2 Comparison with Baselines

To contextualize ClinAgent’s performance, we compare against three baselines: (1) manual programming, (2) generic AI agents without domain tools, and (3) literature-reported automation approaches.

**Key Observations:**

1. **Generic AI agents cannot perform core tasks**: Without ClinAgent tools, agents cannot read SAS datasets or parse Excel specifications—they lack the necessary file format support.
2. **ClinAgent matches manual accuracy for deterministic tasks**: Log analysis and data validation achieve 100% precision, matching or exceeding manual review.
3. **Generative tasks show improvement over generic AI**: Our embedded prompts with domain context achieve 72.1% overall accuracy vs. estimated 30–50% for generic agents without domain guidance.
4. **Effort reduction is substantial**: Preliminary estimates suggest 60–75% reduction in specification and validation time, consistent with Copilot productivity studies [10].

**Limitation**: Direct experimental comparison with generic agents was not performed; generic AI estimates are based on pilot testing and literature. A controlled study comparing ClinAgent-augmented vs. unaugmented agents is future work.

### 7.3 Why Agent-Augmentation Over Custom Agents?

Our architecture choice merits discussion:

By providing tools rather than agents, ClinAgent benefits from rapid improvements in commercial agents (Augment, Claude Code, Cursor) without requiring framework updates.

### 7.4 Implications for Practice

#### 7.4.1 Programmer Role Evolution

ClinAgent tools shift programmer focus:

- **From**: Repetitive coding, manual log review, QC double-programming
- **To**: Specification review, AI output validation, complex logic design

This is augmentation, not replacement—human expertise remains essential for study-specific requirements and edge cases.

**Table 22:** Agent-Augmentation vs. Custom Agent Development.

| Factor | Custom Agent | Agent-Augmentation (ClinAgent) |
| --- | --- | --- |
| Development cost | High (build agent + domain) | Lower (domain tools only) |
| Agent improvements | Manual updates required | Automatic via user’s agent |
| User flexibility | Locked to one system | Use preferred agent |
| Maintenance burden | Full stack | Tools and skills only |
| Enterprise adoption | Change management heavy | Integrates with existing tools |

#### 7.4.2 Quality Assurance Transformation

With deterministic validation tools:

- Log review becomes systematic, not sample-based
- Data validation covers all variables, not spot-checks
- QC can shift from re-creation to verification

#### 7.4.3 Regulatory Pathway

GxP compliance is supported by:

- **Audit trails**: Infrastructure layer logs all operations
- **Deterministic validation**: Rule-based skills produce reproducible results
- **Human oversight**: Skills augment, not replace, human review

### 7.5 Ethical and Regulatory Considerations

The deployment of AI tools in clinical programming raises important ethical and regulatory considerations that ClinAgent’s architecture explicitly addresses.

#### 7.5.1 Data Privacy and PHI Protection

Clinical trial data contains Protected Health Information (PHI) subject to HIPAA, GDPR, and ICH E6(R2) requirements. ClinAgent addresses this through:

- **Local Processing**: All MCP tools run locally; no patient data is transmitted to external AI services unless explicitly configured by the organization.
- **DataMasker Component**: The infrastructure layer includes configurable PHI/PII masking that removes or pseudonymizes sensitive fields before data reaches agent context.
- **Context Minimization**: Skills request only necessary data fields, reducing exposure surface.
- **Audit Logging**: All data access is logged for compliance review.

Organizations retain full control over data handling policies and can configure ClinAgent to meet their specific privacy requirements.

#### 7.5.2 Regulatory Compliance and Validation

FDA 21 CFR Part 11 and EU Annex 11 require validation of computerized systems used in clinical trials. ClinAgent supports compliance through:

- **Deterministic Components**: Rule engines and parsers produce reproducible outputs suitable for validation testing.
- **Audit Trails**: Complete logging of tool invocations, inputs, and outputs.
- **Separation of Concerns**: Validated tools (ClinAgent) are distinct from stochastic components (AI agent reasoning), enabling focused validation scope.
- **Human Oversight**: ClinAgent augments rather than replaces human programmers; all outputs require human review before submission.

We note that regulatory guidance on AI/ML tools in clinical programming is evolving. FDA’s draft guidance on Computer Software Assurance [12] suggests risk-based approaches that ClinAgent’s architecture supports.

#### 7.5.3 Professional Impact and Workforce Considerations

AI automation in clinical programming raises questions about workforce impact. Our position:

- **Augmentation, Not Replacement**: ClinAgent tools assist programmers with repetitive tasks; complex logic, study-specific requirements, and quality judgment remain human responsibilities.
- **Skill Evolution**: As routine tasks become automated, programmer roles shift toward specification review, AI output validation, and complex problem-solving—higher-value activities.
- **Capacity Expansion**: Given industry-wide programmer shortages [2], automation may address capacity constraints rather than displacing workers.

We acknowledge that long-term workforce effects depend on adoption patterns and organizational choices beyond the scope of this technical work.

### 7.6 Limitations

#### 7.6.1 Scope of Validation

Our evaluation validates **tool correctness on one study**. We have not validated:

- End-to-end productivity gains (requires controlled timing studies)
- Generalization across therapeutic areas (requires multi-study evaluation)
- Performance with different AI agents (by design, agent-agnostic)

#### 7.6.2 Ground Truth Dependency

Accuracy metrics depend on expert-created artifacts. If ground truth has errors, our precision/recall calculations may be misleading.

#### 7.6.3 Specification Quality Assumption

Results assume well-structured input specifications. Real-world specifications vary in completeness, and the 4 skipped TLFs (missing macro names) illustrate this dependency.

### 7.7 Future Work

1. **Multi-Study Validation**: Evaluate across Phase 1-3, oncology, CNS, rare diseases
2. **Productivity Studies**: Controlled experiments measuring time savings with ClinAgent tools vs. baseline
3. **Agent Comparison**: Document performance across different AI coding agents
4. **Skill Expansion**: Extend to SDTM mapping, define.xml generation, P21 compliance
5. **Regulatory Engagement**: Early discussions with FDA/EMA on AI tool validation

## 8 Conclusion

### 8.1 Summary

We presented ClinAgent, a skill and tool layer that augments AI coding agents with clinical programming capabilities. Rather than building a custom LLM agent, ClinAgent provides domain-specific tools via the Model Context Protocol (MCP) that any compatible agent can invoke. Our key contributions:

1. **Agent-Augmentation Architecture**: Users keep their preferred AI agent (Augment Code, Claude Code, Cline, Cursor) while gaining clinical programming tools. This enables immediate adoption without workflow disruption and automatic benefit from agent improvements.
2. **“Thin MCP, Thick Skills” Design**: Minimal data access tools (MCP) combined with rich skill packages containing embedded prompts, rule engines, and domain knowledge. This separation enables independent testing and maintenance.
3. **Nine Validated Skills**: SK-001 (Study Setup) through SK-009 (eSub Packaging) covering the clinical programming workflow. Validation using STUDY-A artifacts (specifications, programs, and synthetic data across 11 ADaM domains) confirms:

a. 100% precision on log analysis (1 error, 7 warnings detected)
b. 100% accuracy on data validation (56/56 variables matched)
c. 72.1% derivation accuracy on specification generation
4. **Tool-Centric Evaluation**: Methodology distinguishing tool correctness (our scope) from LLM reasoning performance (agent-dependent, outside our scope).

### 8.2 Practical Implications

ClinAgent’s agent-augmentation approach offers practical advantages:

- **No agent lock-in**: Organizations use their approved AI tools
- **Incremental adoption**: Add clinical tools without replacing workflows
- **Future-proof** : Benefits from agent improvements automatically
- **Testable**: Skills validate independently of agent choice

### 8.3 Future Work

Priority extensions include: (1) multi-study validation across therapeutic areas and phases; (2) controlled productivity studies measuring end-to-end time savings; (3) skill expansion to SDTM, define.xml, and Pinnacle 21 compliance; and (4) regulatory engagement on AI tool validation pathways.

### 8.4 Closing

The question is no longer whether AI can assist clinical programming—modern agents clearly can reason about code. The question is whether domain-specific tools exist to make that reasoning productive. ClinAgent provides those tools, enabling any AI coding agent to become a clinical programming assistant.

The future of clinical programming is human *with* AI: agents provide reasoning, ClinAgent provides domain expertise, and programmers provide oversight and judgment.

## Acknowledgments

The author thanks the clinical programming community for discussions that informed this work, and the developers of the Model Context Protocol for enabling standardized AI tool integration. The author also acknowledges the open-source communities behind the Python libraries used in this research, including pandas, pyreadstat, and openpyxl.

## Ethics Statement

This study involves the development and validation of software tools for clinical trial statistical programming. **No actual patient data was used in this research.** Validation was performed using: (1) study specifications and metadata templates (variable names, data types, derivation logic, and format definitions) from a completed Phase 2 cardiovascular study (referred to as “STUDY-A”); (2) production SAS programs written for that study; and (3) synthetic datasets programmatically generated using the Python Faker library guided by the study specifications. The Faker library [20] generated fictitious values (names, dates, numeric ranges) that conform to specification-defined data types and constraints, producing structurally realistic datasets with-out any connection to real patients. No human subjects were recruited or enrolled for this research. As this research involves only software validation using computationally generated synthetic data, program code, and metadata specifications—none of which contain Protected Health Information (PHI) or personally identifiable information—it does not constitute human subjects research and no IRB approval was required.

## Data Availability Statement

The validation was performed using synthetic datasets, study specifications, and SAS programs—no actual patient data was used. The study specifications and programs from STUDY-A contain proprietary methodology and cannot be shared publicly. The ClinAgent architecture specifications, skill definitions, and example code are available at https://github.com/yanmingyu92/ClinAgent. The validation methodology and detailed results are fully described in this manuscript to enable reproducibility of the approach. Example synthetic datasets matching common ADaM structures are provided in the repository for demonstration purposes.

## Author Contributions

Jaime Yan: Conceptualization, Methodology, Software, Validation, Formal Analysis, Investigation, Data Curation, Writing – Original Draft, Writing – Review & Editing, Visualization.

## A Skill Definitions

### A.1 Complete Skill Specifications

#### A.1.1 SK-001: Study Setup

- **Purpose**: Create standardized folder structure for new study
- **Inputs**: study_id, protocol_number, therapeutic_area
- **Outputs**: Folder hierarchy, template files, configuration
- **Rules**: Naming conventions, required subfolders

#### A.1.2 SK-002: Specification Editor

- **Purpose**: Parse and edit ADaM specifications
- **Inputs**: Excel specification file, edit instructions
- **Outputs**: Updated specification, change log
- **Rules**: Column validation, controlled terminology

#### A.1.3 SK-003: ADaM Coder

- **Purpose**: Generate ADaM SAS programs from specifications
- **Inputs**: Specification, SDTM metadata, macro catalog
- **Outputs**: SAS program file, documentation
- **Rules**: Macro selection, variable derivation patterns

#### A.1.4 SK-004: SAS Runner

- **Purpose**: Execute SAS programs in batch
- **Inputs**: Program path, execution parameters
- **Outputs**: Execution status, log file path, dataset paths
- **Rules**: Environment setup, error handling

#### A.1.5 SK-005: Log Reviewer

- **Purpose**: Analyze SAS logs for issues
- **Inputs**: Log file path, severity threshold
- **Outputs**: Issue list, severity classification, recommendations
- **Rules**: 50+ error patterns, severity classification

#### A.1.6 SK-006: Data Validator

- **Purpose**: Validate datasets against specifications
- **Inputs**: Dataset path, specification, validation rules
- **Outputs**: Validation report, discrepancy list
- **Rules**: Type checking, length validation, controlled terms

#### A.1.7 SK-007: TLF Coder

- **Purpose**: Generate TLF programs from shells
- **Inputs**: TLF shell, ADaM datasets, macro catalog
- **Outputs**: SAS program, mock output
- **Rules**: Macro selection, formatting standards

#### A.1.8 SK-008: RTF QC

- **Purpose**: Compare RTF outputs for QC
- **Inputs**: Production RTF, QC RTF, tolerance settings
- **Outputs**: Comparison report, difference highlights
- **Rules**: Numeric tolerance, whitespace handling

#### A.1.9 SK-009: eSub Packager

- **Purpose**: Create regulatory submission package
- **Inputs**: Study artifacts, submission type
- **Outputs**: eSub folder structure, define.xml
- **Rules**: FDA/EMA requirements, file naming

### A.2 Rule Engine Schema

**Listing 7:**
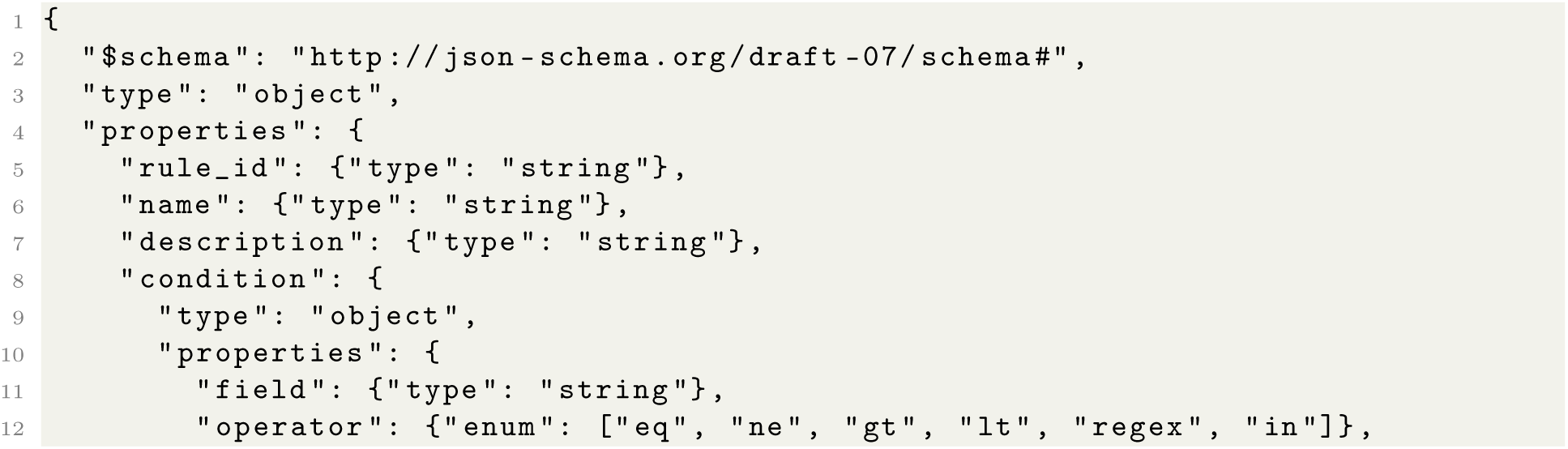
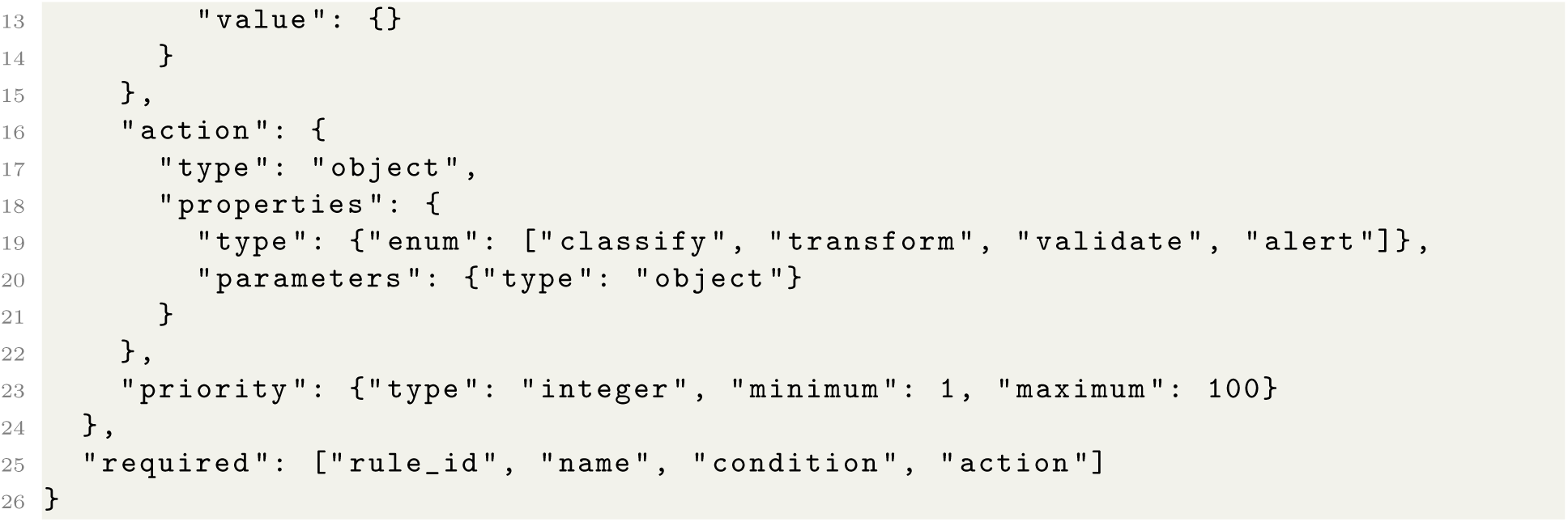
Rule Engine JSON Schema

## B Benchmark Task Specifications

### B.1 ClinProg-Bench-v1 Overview

The benchmark suite comprises 250 tasks designed to evaluate clinical programming automation capabilities.

### B.2 Task Category Specifications

#### B.2.1 T1: Code Generation (60 tasks)

| Subcategory | Description | Count | Complexity |
| --- | --- | --- | --- |
| T1.1 ADaM ADSL | Subject-level dataset | 10 | Mixed |
| T1.2 ADaM BDS | Basic data structure | 20 | Mixed |
| T1.3 ADaM OCCDS | Occurrence data | 10 | Mixed |
| T1.4 TLF Tables | Summary tables | 10 | Mixed |
| T1.5 TLF Listings | Data listings | 5 | Simple |
| T1.6 TLF Figures | Statistical figures | 5 | Complex |

**Acceptance Criteria**:

- Code compiles without errors
- Output matches expected structure
- All specified variables present
- Logic matches specification

#### B.2.2 T2: Code Review (50 tasks)

| Subcategory | Description | Count | Complexity |
| --- | --- | --- | --- |
| T2.1 Log Analysis | Error/warning detection | 20 | Mixed |
| T2.2 Code QC | Logic review | 15 | Mixed |
| T2.3 Style Review | Standards compliance | 15 | Simple |

**Acceptance Criteria**:

- All issues correctly identified
- Severity correctly classified
- No false positives above threshold

#### B.2.3 T3: Specification Interpretation (50 tasks)

| Subcategory | Description | Count | Complexity |
| --- | --- | --- | --- |
| T3.1 Variable Extraction | Parse variable definitions | 20 | Simple |
| T3.2 Derivation Logic | Extract derivation rules | 20 | Mixed |
| T3.3 Ambiguity Detection | Identify unclear specs | 10 | Complex |

#### B.2.4 T4: Documentation (40 tasks)

| Subcategory | Description | Count | Complexity |
| --- | --- | --- | --- |
| T4.1 Program Headers | Generate standard headers | 15 | Simple |
| T4.2 Metadata | Dataset/variable labels | 15 | Simple |
| T4.3 Change Logs | Document modifications | 10 | Simple |

#### B.2.5 T5: Error Diagnosis (50 tasks)

| Subcategory | Description | Count | Complexity |
| --- | --- | --- | --- |
| T5.1 Syntax Errors | Fix compilation errors | 15 | Simple |
| T5.2 Logic Errors | Identify incorrect logic | 20 | Mixed |
| T5.3 Data Issues | Diagnose data problems | 15 | Complex |

### B.3 Protocol

1. **Task Selection**: Random stratified sample from each category
2. **Execution**: Run skill with standardized inputs
3. **Timing**: Record wall-clock time from start to completion
4. **Output Capture**: Store all outputs in standardized format
5. **Expert Review**: Blinded evaluation against gold standard
6. **Scoring**: Apply quality rubric

### B.4 Gold Standard Development

Gold standards were developed by:

- Senior SAS programmers (10+ years experience)
- Independent review by second programmer
- Reconciliation of discrepancies
- Final approval by programming lead

### B.5 Statistical Analysis

- Success rates with 95% Wilson confidence intervals
- Quality scores with bootstrap confidence intervals (1000 resamples)
- Time comparisons with paired t-tests
- Effect sizes with Cohen’s d

